# An integrative analysis of consortium-based multi-omics QTL and genome-wide association study data uncovers new biomarkers for lung cancer

**DOI:** 10.1101/2024.12.13.24318992

**Authors:** Yanru Wang, Aoxuan Wang, Ning Xie, Xiaowen Xu, Xiang Wang, Mengshen Zhao, Xuan Wang, Jiacheng Zhou, Yang Zhao, Zhibin Hu, Hongbing Shen, Rayjean J. Hung, Christopher I. Amos, Yi Li, David C. Christiani, Feng Chen, Yongyue Wei, Ruyang Zhang

**Affiliations:** Department of Biostatistics, Center for Global Health, School of Public Health, Nanjing Medical University, Nanjing, Jiangsu 211166, China; China International Cooperation Center (CICC) for Environment and Human Health, Nanjing Medical University, Nanjing, Jiangsu 211166, China; Department of Epidemiology, School of Public Health, Nanjing Medical University, Nanjing, Jiangsu 211166, China; Jiangsu Key Lab of Cancer Biomarkers, Prevention and Treatment, Cancer Center, Collaborative Innovation Center for Cancer Personalized Medicine, Nanjing Medical University, Nanjing, Jiangsu 211166, China; State Key Laboratory of Reproductive Medicine, Nanjing Medical University, Nanjing, Jiangsu 211166, China; Lunenfeld-Tanenbaum Research Institute, Sinai Health, and University of Toronto, Toronto, ON, Canada M5G 1X5; The Institute for Clinical and Translational Research, Baylor College of Medicine, Houston, TX 77030, USA; Department of Biostatistics, University of Michigan, Ann Arbor, MI 48109, USA; Department of Environmental Health, Harvard T.H. Chan School of Public Health, Boston, MA 02115, USA; Pulmonary and Critical Care Division, Department of Medicine, Massachusetts General Hospital and Harvard Medical School, Boston, MA 02114, USA; Center for Public Health and Epidemic Preparedness & Response, Peking University, Beijing, 100191, China; Key Laboratory of Epidemiology of Major Diseases (Peking University), Ministry of Education, Beijing, 100191, China; Changzhou Medical Center, Nanjing Medical University, Changzhou, Jiangsu 213164, China; Information Center, The Affiliated Changzhou Second People’s Hospital of Nanjing Medical University, Changzhou, Jiangsu 213164, China

## Abstract

The role of molecular traits (e.g., gene expression and protein abundance) in the occurrence, development, and prognosis of lung cancer has been extensively studied. However, biomarkers in other molecular layers and connections among various molecular traits that influence lung cancer risk remain largely underexplored. We conducted the first comprehensive assessment of the associations between molecular biomarkers (i.e., DNA methylation, gene expression, protein and metabolite) and lung cancer risk through epigenome-wide association study (EWAS), transcriptome-wide association study (TWAS), proteome-wide association study (PWAS) and metabolome-wide association study (MWAS), and then we synthesized all omics layers to reveal potential regulatory mechanisms across layers. Our analysis identified 61 CpG sites, 62 genes, 6 proteins, and 5 metabolites, yielding 123 novel biomarkers. These biomarkers highlighted 90 relevant genes for lung cancer, 83 among them were first established in our study. Multi-omics integrative analysis revealed 12 of these genes overlapped across omics layers, suggesting cross-omics interactions. Moreover, we identified 106 potential cross-layer regulatory pathways, indicating that cell proliferation, differentiation, immunity, and protein-catalyzed metabolite reaction interact to influence lung cancer risk. Further subgroup analyses revealed that biomarker distributions differ across patient subgroups. To share all signals in different omics layers with community, we released a free online platform, LungCancer-xWAS, which can be accessed at http://bigdata.njmu.edu.cn/LungCancer-xWAS/. Our findings underscore the importance of xWAS which integrating various types of molecular quantitative trait loci (xQTL) data with genome-wide association study (GWAS) data to deepen understanding of lung cancer pathophysiology, which may provide valuable insights into potential therapeutic targets for the disease.

## Main

Lung cancer, the most frequently diagnosed cancer representing roughly one in eight cases globally in 2022, remains the leading cause of cancer-related deaths, accounting for 18% of all cancer fatalities worldwide^1^. As a complex disease, lung cancer is driven by a combination of various environmental and genetic factors^2,3^. Over the past decade, genome-wide association studies (GWASs) have identified more than 50 genetic markers for lung cancer risk^4–6^. However, the underlying mechanism remains unclear, limiting a deeper understanding of the disease and the translation of these genetic findings into clinical practice^7^.

Recent advances in high-throughput omics platforms have greatly increased the availability of various types of molecular quantitative trait loci (xQTL) summary statistics, including meQTL (DNA methylation), eQTL (gene expression), pQTL (protein) and metabQTL (metabolite)^8^. Researchers have since proposed xWAS frameworks^9–11^, which integrate GWAS data with xQTL data to uncover potential mechanisms and identify candidate therapeutic targets beyond those derived from genetic studies^7^. By leveraging a reference panel of genotypes and molecular traits, xWAS frameworks model regulations of different levels of genetic-determinant molecular traits and enable the estimation of biomolecule-disease associations without using individual-level omics data^9^. Currently, these frameworks have been successfully applied to a variety of phenotypes, including neurological and psychiatric phenotypes^11^, autoimmune diseases^9^ and osteoarthritis^12^.

Previous studies have reported the associations of molecular traits on lung cancer using the xWAS frameworks, including epigenome-wide association study (EWAS), transcriptome-wide association study (TWAS) and proteome-wide association study (PWAS) ^10,13–16^. For example, an EWAS identified associations between 16 genetically predicted CpGs and non–small cell lung cancer (NSCLC)^16^, and a TWAS discovered a novel lung adenocarcinoma (LUAD) susceptibility gene whose expression predominantly driven by a locus on 9p13.3^13^. However, existing studies have relied on a single type of xQTL dataset with relatively small sample sizes, making it challenging to interpret GWAS signals with respect to general populations as well as specific subgroups.

In this study, we aimed to identify molecular traits and potential regulatory pathways for lung cancer by integrating alliance-based xQTL summary statistics from multiple omics layers with consortium-based lung cancer GWAS data. To detect lung cancer related biomarkers, we conducted xWAS analyses, including TWAS, EWAS, PWAS, and metabolome-wide association study (MWAS), across overall and subgroup populations. We then synthesized all xWAS association results to nominate candidate genes for lung cancer across different omics layers. Additionally, we systematically analyzed correlations among molecular traits from different omics layers, focusing on pathways with consistent association directions to explore potential regulatory mechanisms.

## Results

We conducted xWAS of lung cancer by integrating xQTL summary-level statistics from 4 omics layers, including DNA methylation markers (*n* = 244,533)^17^, gene expressions (*n* = 19,942)^18^, proteins (*n* = 4,907)^19^ and metabolites (*n* = 486)^20^ from blood, along with individual-level lung cancer GWAS datasets sourced from The International Lung Cancer OncoArray Consortium (ILCCO-OncoArray, 16,606 cases and 13,905 controls), The Transdisciplinary Research In Cancer of the Lung (TRICL, 4,975 cases and 5,484 controls), The Prostate, Lung, Colorectal, and Ovarian) (PLCO, 1,787 cases and 85,821 non-cases) and UK Biobank (UKB, 4,369 cases and 344,700 non-cases) (Methods and Supplementary Table 1). In this study, associations between lung cancer and genetically predicted CpGs, gene expressions, proteins and metabolites were identified through a two-phase (discovery and replication) analytic strategy, complemented by meta-analyses, multi-phase association analyses, sensitivity analyses and subgroup analyses (Fig.1). Furthermore, we synthesized all association test results to nominate genes and reveal those overlapped across different omics layers. Building on these results, we explored potential regulatory pathways where significant genes exhibiting correlations across multiple omics layers.

**Fig. 1.**
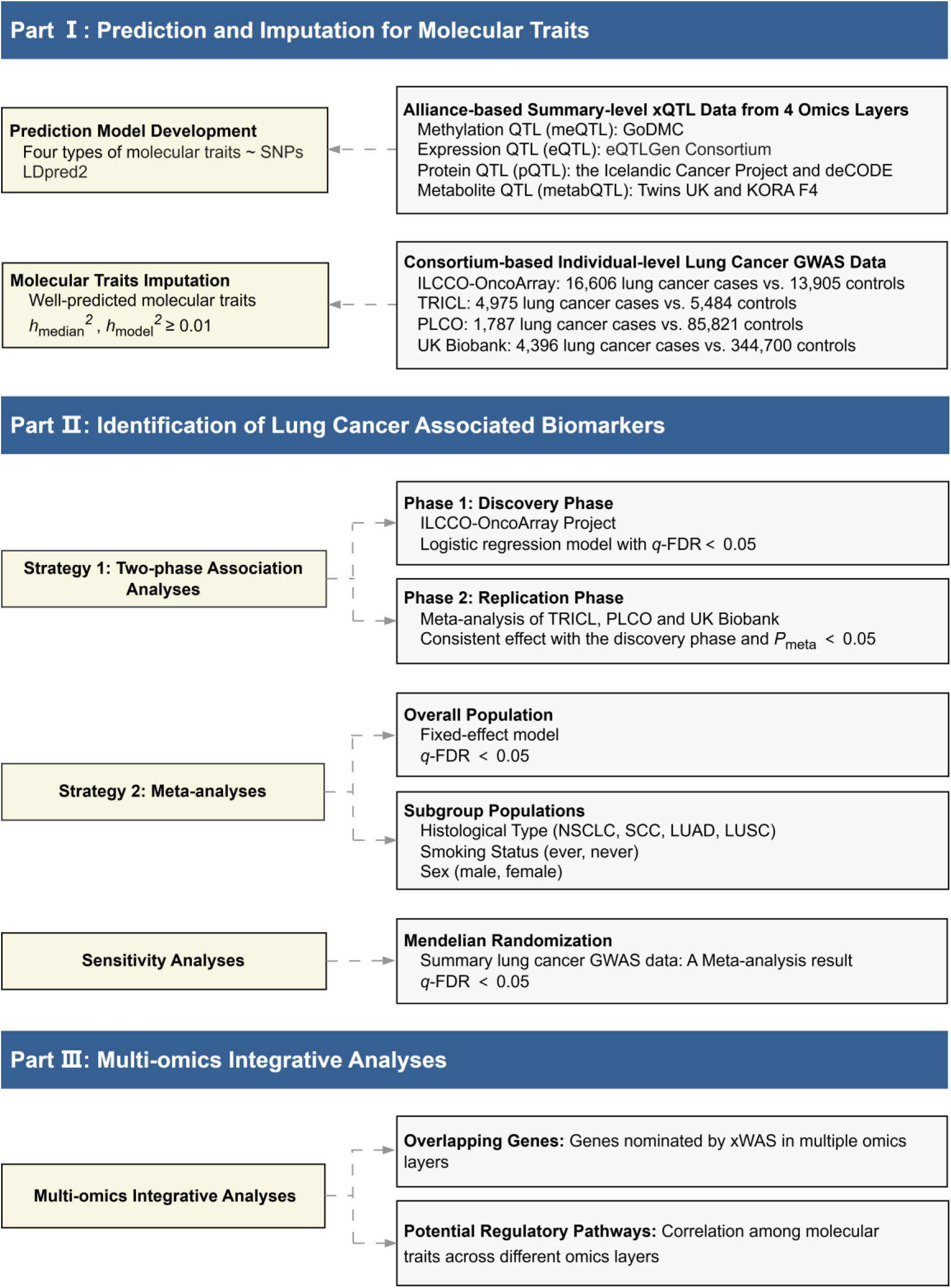
Flowchart for the study design.

### Development of Molecular Traits Prediction Models

We used LDpred2 to predict molecular traits based on alliance-based xQTL summary-level statistics. In total, 145,585 molecular traits were successfully predicted, including 127,497 (52.1%) CpGs, 13,039 (65.4%) gene expressions, 4,410 (89.9%) proteins and 320 (65.8%) metabolites. We used heritability values *h*^2^_median_ and *h*^2^_model_ to evaluate the predictive ability of SNPs. Large variation was observed in *h*^2^_median_, *h*^2^_model_, and the number of variants included in the prediction models across all layers (Supplementary Fig. 1; Supplementary Table 2-5). According to the commonly used threshold of heritability 0.01, among these successfully predicted molecular traits, 89,073 (69.9%) DNA methylation models, 9,012 (69.1%) expression models, 4,225 (95.8%) protein models, and 140 (43.8%) metabolite models, which possessed a satisfactory predictive performance (*h*^2^_median_ ≥ 0.01 and *h*^2^_model_ ≥ 0.01), were retained for the subsequent analyses (Supplementary Fig. 1).

### Novel Biomarkers Associated with Lung Cancer Identified by Two-phase Association Analyses

#### EWAS

In the discovery phase, 66 CpGs reached the significance threshold after false discovery rate (FDR) correction (*q*-FDR < 0.05) in ILCCO-OncoArray, and 61 significant CpGs with the consistent effect direction and *P*_meta_ < 0.05 were confirmed in the replication phase (Fig. 2A). In general, CpG markers were mainly located in the well recognized lung cancer susceptibility regions, including 5p15.33 and 15q25.1. Of these 61 CpGs, 54 are reported for the first time in our study, while the remaining 7 had been previously reported^16,21^ (Supplementary Table 6, 22). Please see Methods section for the definition of novel biomarkers and genes. A positive association of 25 CpGs with lung cancer risk was detected, whereas the other 36 CpGs were negatively associated with lung cancer (Supplementary Table 6). Specifically, cg20622131 (at 15q25.1) was the risk biomarker with the largest effect size (discovery phase: *OR* = 2.18, 95%CI: 1.84-2.59, *P* = 9.26×10^-19^, *q*-FDR = 7.50×10^-15^; replication phase: *OR* = 2.27, 95%CI: 1.92-2.67, *P* = 3.91×10^-22^) and cg20711912 (at 5p15.33) had the largest protective effect (discovery phase: *OR* = 0.34, 95%CI: 0.26-0.43, *P* = 1.72×10^-19^, *q*-FDR = 1.91×10^-15^; replication phase: *OR* = 0.35, 95%CI: 0.28-0.44, *P* = 1.13× 10^-20^).

**Fig. 2.**
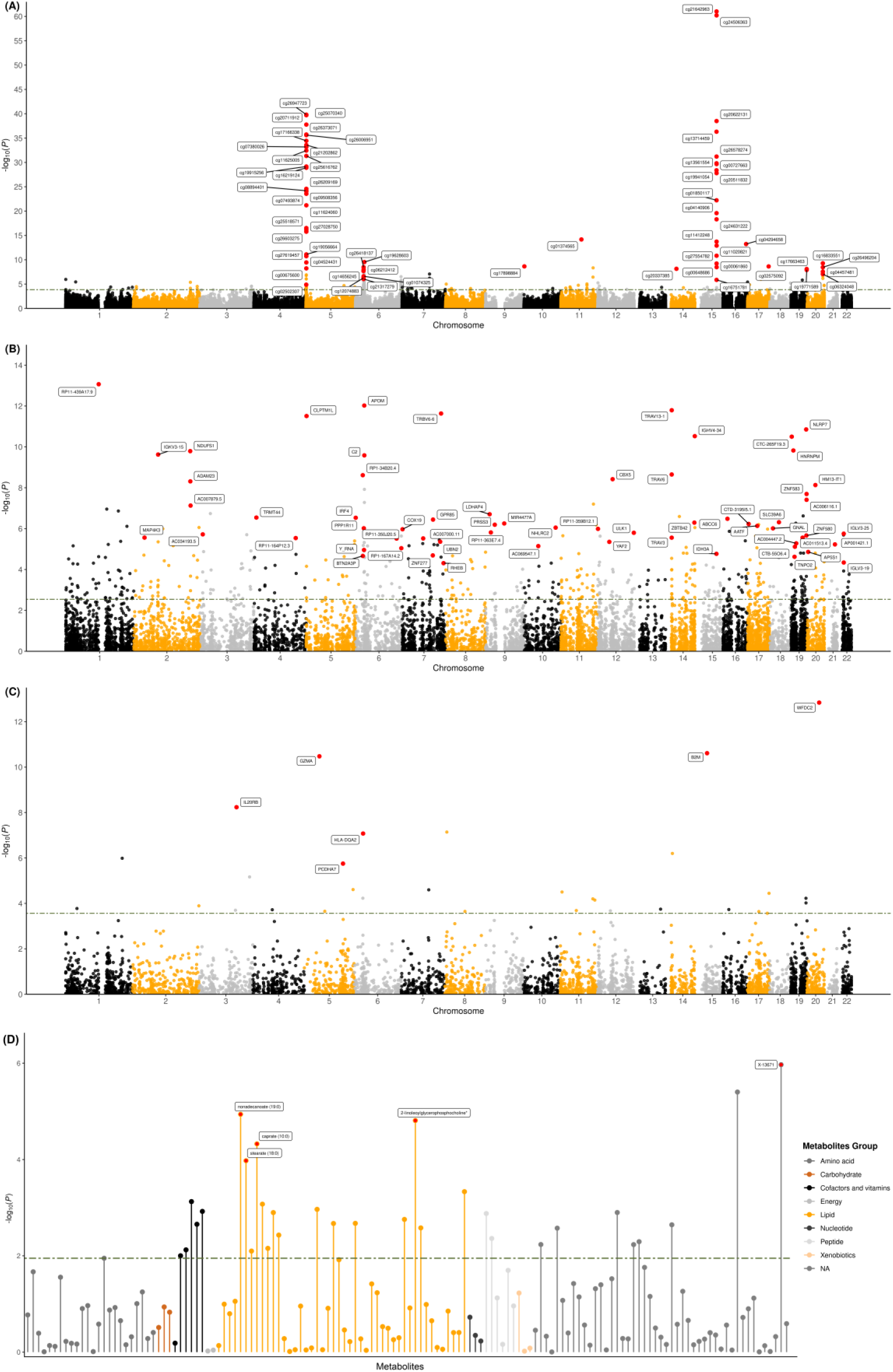
Results of xWAS for the associations between genetically predicted molecular traits and lung cancer in four omics layers. (A) Manhattan plot for DNA methylation analyses; (B) Manhattan plot for gene expression analyses; (C) Manhattan plot for protein analyses; (D) Lollipop plot for metabolite analyses. The green dotted line reflects the significant threshold of the *q*-FDR < 0.05. Each dot represents the genetically predicted molecular trait. The x axis represents the chromosome of the molecular traits in Manhattan plots of methylation, gene expression and protein, and represents metabolite in Lollipop plot of metabolite. The y axis represents the negative logarithm of the association *P* value.

#### TWAS

In the discovery phase, 91 genes reached the significance threshold (*q*-FDR < 0.05), and 62 genes were successfully validated with *P*_meta_ < 0.05 in the replication phase (Fig. 2B). Of these 62 genes, 60 are newly identified to be associated with lung cancer by our study (Methods), while 2 (*APOM* and *CLPTM1L*) were reported before^22^ (Supplementary Table 7, 22). A positive association with lung cancer risk was found for 32 genes, while the remaining 30 genes were negatively associated with lung cancer (Supplementary Table 7). Specifically, *RP11-350J20.5* was the risk gene with the largest effect size (discovery phase: *OR* = 3.69, 95%CI: 1.93-7.07, *P* = 7.80×10^-5^, *q*-FDR = 1.67×10^-2^; replication phase: *OR* = 2.33, 95%CI: 1.26-4.34, *P* = 7.38×10^-3^) and *HM13-IT1* was the gene with the highest protective effect (discovery phase: *OR* = 0.29, 95%CI: 0.16-0.52, *P* = 3.22×10^-5^, *q*-FDR = 9.86×10^-3^; replication phase: *OR* = 0.33, 95%CI: 0.19-0.57, *P* = 5.53×10^-5^).

#### PWAS

In the discovery phase, 7 proteins reached the significance threshold (*q*-FDR < 0.05), while 6 proteins with the consistent effect direction and *P*_meta_ < 0.05 were identified in the replication phase (Fig. 2C). Of them, 4 (WAP four-disulfide core domain protein 2, granzyme A, beta-2-microglobulin and protocadherin alpha-7) are novel proteins associated with lung cancer (Methods), while 2 (interleukin-20 receptor subunit beta and HLA class II histocompatibility antigen) had been reported previously (Supplementary Table 8, 22).

#### MWAS

In the discovery phase, 15 significant (*q*-FDR < 0.05) metabolites were discovered, and 5 novel metabolites [2-linoleoylglycerophosphocholine, stearate (18:0), nonadecanoate (19:0), caprate (10:0) and an unknown metabolite coded X-13671] were certified as robust (*P*_meta_ < 0.05) biomarkers in the replication phase (Fig. 2D and Supplementary Table 9). Among them, four known metabolites were classified as lipids, and all showed a negative association with lung cancer.

The quantile-quantile (Q-Q) plots for the xWAS in both two phases, along with heatmaps depicting the correlation between molecular traits in each layer, are provided in Supplementary Fig. 2-5. Additionally, significance status of these biomarkers in each dataset (i.e., OncoArray, TRICL, PLCO and UKB) were summarized in Supplementary Tables 10-13.

### Sensitivity Analyses

To validate whether the xWAS associations could be replicated using an alternative methodology, a two-sample Mendelian Randomization (MR) analysis was conducted for each biomarker-lung cancer association. Thirty-four associations with the same direction as estimated by xWAS remained significant after FDR correction (*q*-FDR < 0.05), including 15 CpGs, 15 genes, 2 proteins, and 2 metabolites (Supplementary Table 14-17).

### Subgroup-specific Signals Identified by Meta-Analyses

We conducted subgroup analyses by histological type [NSCLC, squamous cell carcinoma (SCC), LUAD and lung squamous cell carcinoma (LUSC)], smoking status [ever (current and former) smokers and never smokers] and gender. Lung cancer associated biomarkers with *q*-FDR < 0.05 derived from meta-analyses of four datasets in overall and subgroup populations (Methods) were exhibited in Supplementary Fig. 6-13 and summarized in Supplementary Table 18-21. Signals varied markedly among the various subpopulations.

#### EWAS

Overall, 499 CpG markers reached the significance threshold (*q*-FDR < 0.05) in different subgroups (Supplementary Table 18). CpG markers of NSCLC participants mainly located in 5p15.33, 6q27 and 15q25.1 regions (Supplementary Fig. 6A). These signals for SCC patients overlapped with those for NSCLC in region 15q25.1 (Supplementary Fig. 7A). The 5p15.33 region was abundant in shared methylation sites between smokers and non-smokers, whereas CpG markers in 6p21.32, 6p21.33, 6p22.1, 6q27, 7q22.1 and 15q25.1 regions were significant only in smokers (Supplementary Fig. 8A, 9). In male and female subgroups, the signals were mainly located in the same regions, such as 5p15.33 and 15q25.1 (Supplementary Fig. 10A, 11A).

#### TWAS

Totally, 930 genes with *q*-FDR < 0.05 were observed in different subgroups (Supplementary Table 19; Supplementary Fig. 6B, 7B, 8B, 10B, 11B). No significant associations were found in non-smokers, while 29 NSCLC-specific genes, 25 SCC-specific genes, 235 smoker-specific genes, 33 male-specific genes, and 4 female-specific genes were identified, respectively. Interestingly, *IGKV3-15* was significant in all subgroups, which was positively associated with lung cancer risk.

#### PWAS

We discovered 79 proteins with *q*-FDR < 0.05 in different subgroups (Supplementary Table 20; Supplementary Fig. 6C, 8C, 10C). Of these, 28 proteins showed subgroup-specific associations, including 5 in NSCLC participants and 23 in smokers. Two proteins (WAP four-disulfide core domain protein 2, Granzyme A) were identified associated with lung cancer with the same effect direction in all groups: protein with a positive effect was WAP four-disulfide core domain protein 2 and Granzyme A were negatively associated with lung cancer. However, no significant associations were found in SCC, female participants, or non-smokers.

#### MWAS

We observed 36 metabolites with *q*-FDR < 0.05 in different subgroups (Supplementary Table 21; Supplementary Fig. 6D, 8D, 10D, 11C). No significant associations were found in SCC participants and non-smokers. However, 3 subpopulation-specific metabolites were identified, including 1 in female participants (2-aminobutyrate) and 2 in smokers (1-palmitoylglycerophosphoethanolamine and X-11315). Nonadecanoate (19:0) was a risk factor across all groups.

To share all signals in different omics layers with community, we released a free online platform, LungCancer-xWAS, which can be accessed at http://bigdata.njmu.edu.cn/LungCancer-xWAS/.

### Nominating Overlapping Genes Across Different Omics Layers

Totally, we identified 134 significant molecular biomarkers associated with lung cancer in xWAS through the two-phase analytic strategy. These biomarkers were mapped into 90 genes based on their physical positions (Fig. 3), including novel 83 genes that never been previously reported (Methods and Supplementary Table 22). We also revealed genes overlapped across different omics layers, i.e. genomics layer, epigenomics layer, transcriptomics layer, proteomics layer and metabolomic layer. Genes were nominated as overlapping genes if meeting the following criteria: (1) they were nominated as significant genes by the two-phase analytic strategy in any one of layers or by previous GWASs (Methods), and (2) they showed nominal significance in the meta-analysis of all datasets (*P* < 0.05) in at least one of other omics layers. Based on this strategy, there were 4 genes overlapping across the genomics layer and epigenomics layer (*TERT*, *CHRNA5*, *CHRNA3* and *CHRNB4*), 5 genes overlapping across the epigenomics layer and transcriptomics layer (*PPP1R2P1*, *RHEB*, *SLC39A6*, *ZNF583*, and *COX19)*, 2 genes overlapping across the transcriptomics layer and proteomics layer (*ABCC6*, *C2*), and gene *CLPTM1L* overlapping across the genomics layer, epigenomics layer and transcriptomics layer (Fig. 3). These overlapping genes suggest that the corresponding molecular traits may be involved in the same regulatory pathway of lung cancer.

**Fig. 3.**
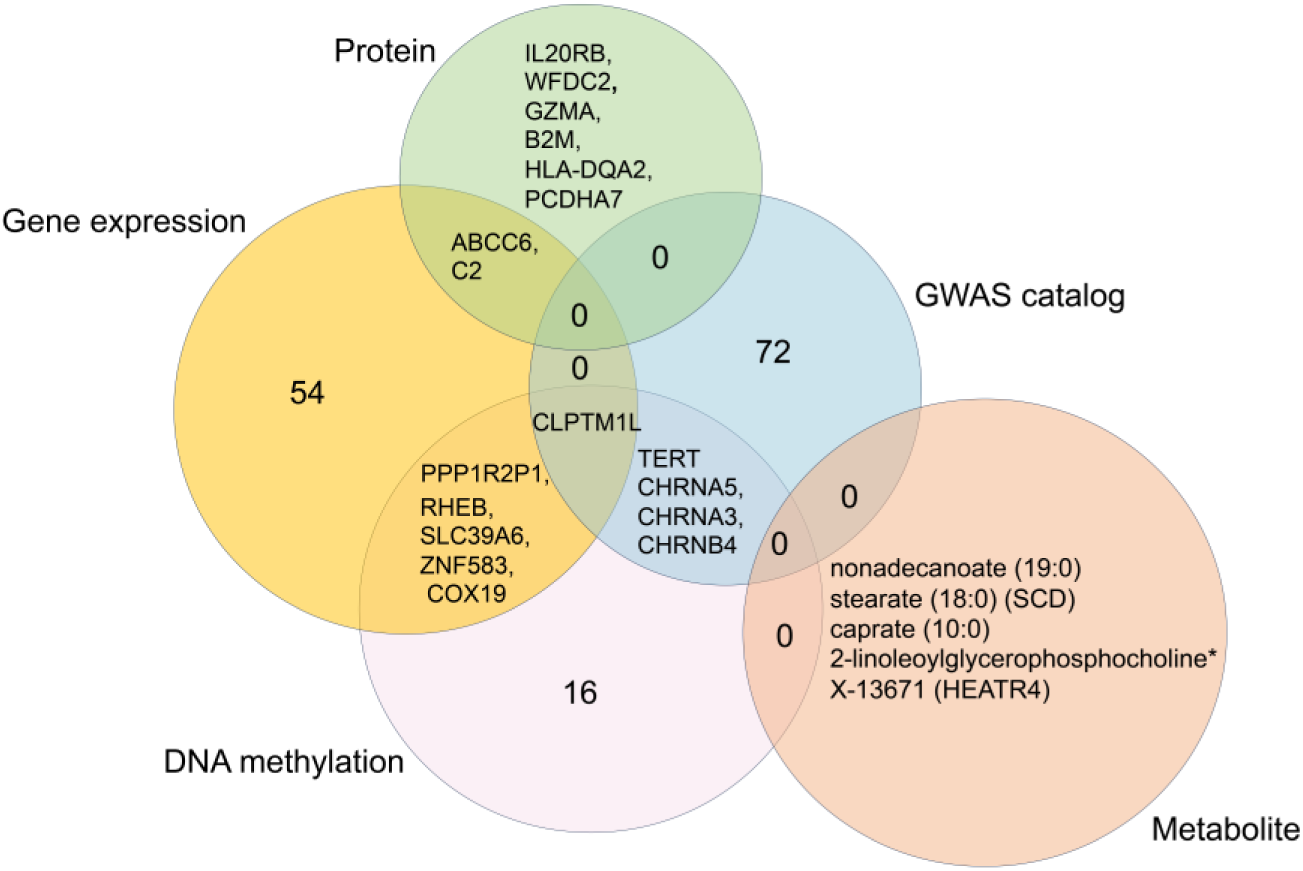
Genes nominated and overlapped across different omics layers by xWAS. For genes nominated less than six by a single type of molecular trait, the prioritized genes or biomarkers instead of the number were listed.

### Exploring Potential Cross-layer Pathways through Multi-omics Integrative Analyses

By assessing the correlation between biomarkers identified by xWAS, we aim to uncover the underlying molecular mechanisms of lung cancer explained by these cross-omics relationships (Methods). We first evaluated the correlations between molecular traits having overlapping nominated genes. We observed a series of pathways for 6 overlapping genes. Seventeen CpGs and 5 genes were involved in the DNA methylation-expression-lung cancer regulatory pathways, one of which has been previously reported^16^. In addition, we discovered one expression-protein-lung cancer regulatory pathway (Fig. 4A; Supplementary Table 23). For example, with cg20711912 and the gene expression level of *CLPTM1L* sharing the overlapping gene *CLPTM1L*, we examined the correlation between cg20711912 and *CLPTM1L*. The methylation marker cg20711912, which is a protective factor of lung cancer, was positively associated with expression of *CLPTM1L* (*r* = 0.45, *q*-FDR < 0.0001). Meanwhile, *CLPTM1L* expression may exert a protective effect on lung cancer via regulating DNA methylation cg20711912.

**Fig. 4.**
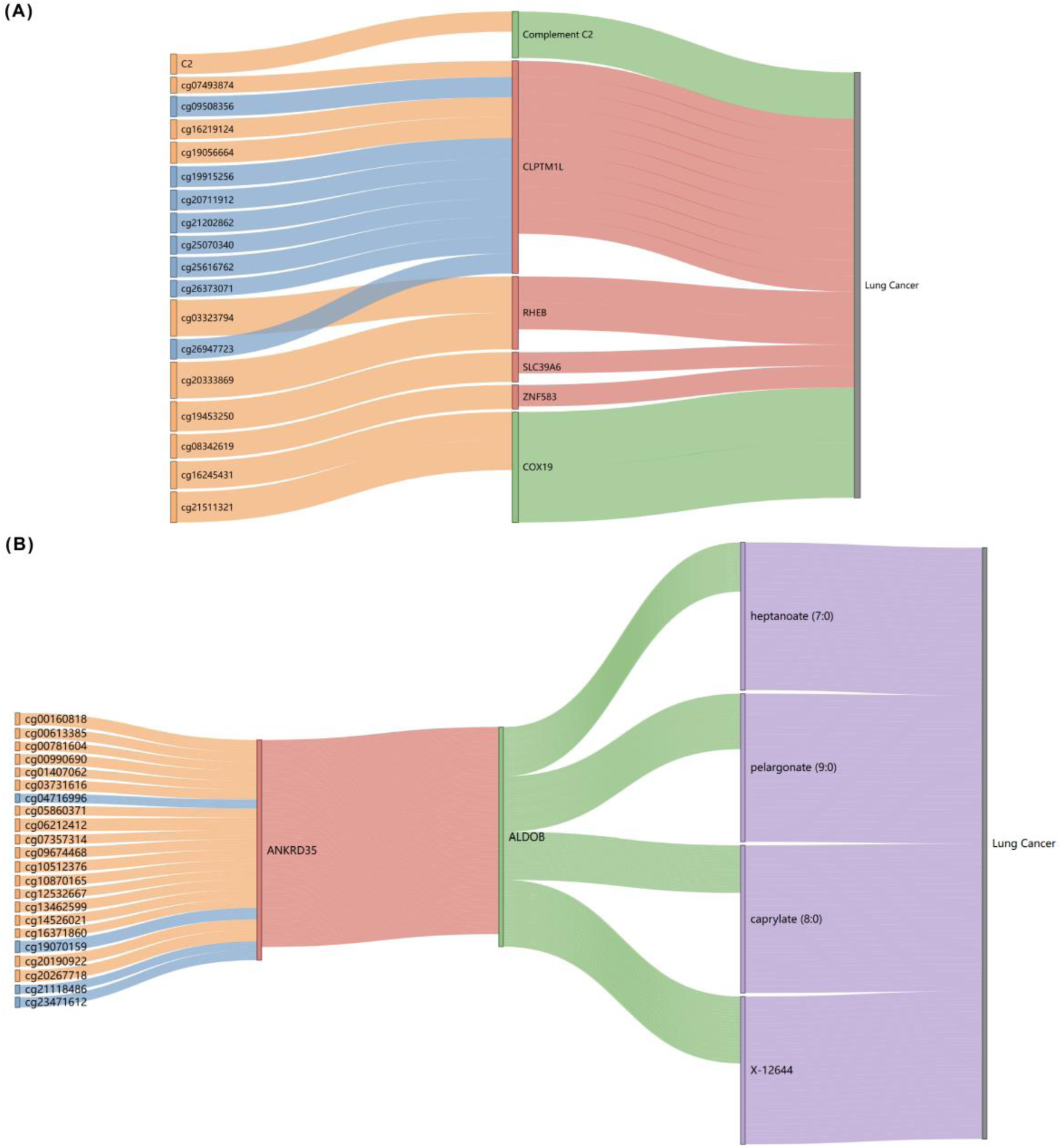
Potential across-layer regulatory pathways for lung cancer. (A) Potential cross-layer pathways for biomarkers mapping to the same overlapping genes. The blue of the left and the red of the middle represent those biomarkers were negatively associated with lung cancer, and the orange of the left and the green of the middle represent those biomarkers were positively associated with lung cancer. The top line is the gene expression-protein-lung cancer pathway, and others were DNA methylation-gene expression-lung cancer pathways. (B) Potential cross-layer pathways consisting of multiple genes with biomarkers including CpGs, gene expressions, proteins and metabolites. The blue of CpG markers represents negatively associated with lung cancer and the orange represents positively associated with lung cancer.

Similarly, by checking correlation among molecular traits across all layers, we found 88 potential regulatory pathways of lung cancer by a hill-climbing strategy (Fig. 4B; Supplementary Table 24). For example, DNA methylation cg06212412 (*TRIM26*) associated with an increased lung cancer risk was negatively (*r* = -0.45, *q*-FDR < 0.0001) correlated with gene expression of *ANKRD35*, which was a protective factor of lung cancer and was positively (*r* = 0.33, *q*-FDR < 0.0001) associated with protein fructose-bisphosphate aldolase B, which was a protective factor of lung cancer. Meanwhile, the protein fructose-bisphosphate aldolase B was positively (*r* = 0.32, *q*-FDR < 0.0001) associated with metabolite heptanoate (7:0), which was a protective factor of lung cancer. That is, cg06212412 may affect the development of lung cancer through regulating *ANKRD35* expression, protein fructose-bisphosphate aldolase B and then metabolite heptanoate (7:0).

## Discussion

In this study, we conducted a comprehensive multi-omics integrative analysis within xWAS framework by leveraging four large-scale lung cancer consortium-based individual-level lung cancer GWAS datasets and alliance-based xQTL summary-level data. We identified 123 novel biomarkers and nominated 83 novel genes for lung cancer. Besides, 12 overlapping genes across different omics layers and 106 potential cross-layer potential molecular pathways were discovered. These findings highlighted the critical role of integrating xQTL data from multiple omics layers with GWAS data in studying the molecular mechanisms of complex diseases.

The first step of xWAS was to develop prediction models of molecular traits. The SNPs that reached the genome-wide significance level with the Bonferroni correction (5× 10^-8^ = 0.05/the number of biomarkers tested) were included in the development of the prediction model using LDpred2. Compared to previous studies, the median *h*^2^_model_ was lower in our MWAS (0.010) and relatively consistent in EWAS (0.023), TWAS (0.025), and PWAS (0.062)^9,16,23^. Since metabolites are downstream products, the genetic-determinant proportion might be less than the upstream molecular traits, such as DNA methylation or gene expression. And there are challenges of using the metabQTL summary-level data rather than raw individual-level data, and the use of the entire genome for prediction rather than just *cis*-SNPs or significant signals. Nonetheless, among the 320 metabolites genetically predicted by LDpred2, 140 (43.75%) metabolites could still be well predicted at or above the *h*^2^_median_ and the *h*^2^_model_ threshold of 0.01, which was commonly used by previous studies to filter out poorly predicted biomolecules. This supports the feasibility of our MWAS performing comparably to EWAS, TWAS and PWAS in testing associations.

Previous studies have observed an 8% improvement in lung cancer discrimination by adding 6 CpGs into conventional risk prediction models^24^. Similarly, an early screening and diagnostic models for NSCLC that integrated proteins and metabolites outperformed classical clinical panel alone^25^. These evidences hint that integrating biomarkers across multiple omics layers from our xWAS will lead to the development of higher performing lung cancer risk prediction and diagnostic models. Our xWAS identified 134 significant biomarkers across different layers, which enables a chance for improvement of lung cancer prediction model. Among them, 54 CpG sites, 60 gene expressions, 4 proteins and 5 metabolites have not been reported in previous publications^10,14,15,26–31^ Our EWAS identified novel lung cancer associated DNA methylation regions, such as cg19628603 in 6p21.32, cg19771589 and cg17663463 in 19q13.43. Besides, cg11412248 (15q25.1) was associated with follicular thyroid carcinoma^32^ and papillary thyroid carcinoma^32,33^, and cg26496204 (20q13.32) in novel regions was associated with B acute lymphoblastic leukemia^34^, indicating a potential methylation phenomenon of multicancer risk. Our TWAS identified several novel genes for lung cancer not reported before, such as *ZNF277* in 7q31.1. *ZNF277* represses transcription of the tumor suppressor gene *INK4A*/*ARF* (recorded in GeneCards). Granzyme A (GZMA), the protein with the highest protective effect, was first identified associated with lung cancer by our PWAS. As a T cell- and natural killer cell-specific serine protease, GZMA can catalyze cleavage of gasdermin-B gasdermin-B (GSDMB), releasing its pore-forming N-terminal domain, which provokes cancer cell death^35^. The majority of metabolites identified by our MWAS were fatty acids (FAs), including nonadecanoate (19:0), stearate (18:0) and caprate (10:0). These were negatively associated with lung cancer due to their roles in membrane formation, energy storage, and tumor signaling^36^. Decreased levels of FAs, such as caprate (10:0), nonadecanoate (19:0), and stearate (18:0), have also been observed in the plasma of East Asian lung cancer patients^37^, which can support our results.

GWAS have identified various genes associated with lung cancer, but the underlying mechanisms for lung cancer remain unclear^4–6^. Our xWAS showed that integrating xQTL summary-level data from multiple layers with lung cancer GWAS data not only recovered findings of previous studies, but also nominated a series of novel genes for lung cancer. These genes have multiple potential biological functions relevant of lung cancer. For example, gene *SCD* located in 10q24.31 was first nominated for lung cancer by our MWAS in genetically determined metabolite level for European ancestry, which encodes an enzyme involved in fatty acid biosynthesis and plays a key role in the double bound into palmitic and stearic acids. It has been recently demonstrated that *SCD* is highly expressed in LUAD from Chinese patients and promotes in vitro and in vivo tumorigenesis, cell migration and invasion^38^. We explored genes nominated by xWAS overlapped across different omics layers by integrating all association test results. We identified 12 genes overlapping across different omics layers, and for those that did not overlap they often participant in different cellular functions and contribute to unrelated organismal traits^8^. Gene *CLPTM1L* located in 5p15.33, one of the well-known susceptibility genes for lung cancer, was overlapped across the genomics layer, epigenomics layer and transcriptomics layer. Besides, the other overlapping genes are also emerged. For example, gene *PPP1R2P1* located in 6p21.32, overlapped across the epigenomics layer and transcriptomics layer, encodes a phosphatase inhibitor. Regulatory T cells (Tregs) are immunosuppressive, and their increased presence can promote tumor growth and progression^39,40^. Recent evidence has illustrated the increase in the number of Treg cells in the tumor microenvironment^40^. Silencing *PPP1R11* has been discovered to make T cells resistant to Treg-mediated suppression of TCR-induced cytokine expression, facilitating cancer immunotherapy^41^. Gene *C2* is located in 6p21.33, whose increased expression level increases the risk of lung cancer-related respiratory diseases, such as asthma^42,43^. Therefore, it is reasonable to hypothesize that genes like *C2* may affect respiratory system diseases, thereby indirectly promoting the initiation and growth of lung cancer. Additionally, the gene *CHRNA5* on 15q25.1, which encodes a nicotinic acetylcholine receptor subunit, overlapped across the genomics and epigenomics layers and is positively associated with lung cancer ^6^.

Interestingly, our subgroup analyses revealed a significant association heterogeneity between smokers and nonsmokers at *CHRNA5* methylation sites (cg01850117, cg16751781, cg21642963 and cg24631222). Previous studies detected an upregulation of the *CHRNA5* gene in NSCLC tumor tissue^44,45^. Still, some researchers also found that lower expression of *CHRNA5* was causally associated with an increased risk of lung cancer through TWAS framework and MR analysis^13,46^. Among 4 significant *CHRNA5* DNA methylation sites in smokers, 3 were protective factors and the other one was a risk factor. Therefore, we hypothesized that these CpG sites may affect lung cancer indirectly through different molecular pathways.

By integrating all association test results from xWAS, we identified pathways with consistent association directions, which could help elucidate the potential regulatory mechanisms involved. First, we examined correlations between molecular traits with overlapping genes. Take gene *RHEB* as an example. It locates in 7q36.1, overlapped across the epigenomics layer and transcriptomics layer, which is a member of the Ras superfamily of small GTPases. *RHEB* signaling drives tumorigenesis and affects apoptosis, cellular senescence, and treatment responses by stimulating mTORC1 (mechanistic/mammalian target of rapamycin complex 1)^47^. Researchers proposed that farnesyl transferase inhibitors (FTIs) could be useful in treating tumors associated with Rheb/mTORC1 activation, considering RHEB needs to be farnesylated to activate mTORC1^48^. Moreover, FTIs inhibit RHEB/mTORC1 activity in NSCLC cell lines ^49^. Therefore, RHEB exhibits a predictive and prognostic significance and may be a promising therapeutic target of lung cancer. We found that DNA methylation sites of *RHEB* are positively associated with lung cancer and may influence the disease by regulating *RHEB* expression. Similarly, DNA methylation of *SLC39A6*, located on 18q12.2, is positively associated with lung cancer and may affect the disease through regulation of *SLC39A6* expression. *SLC39A6* is a member of the zinc transporter subfamily, playing a crucial role in regulating intracellular zinc homeostasis^49,50^. Zinc usually has an important role in tumor events, and it has been confirmed that lung cancer patients presented with significantly lower levels of zinc compared to controls^51,52^.

Second, we examined the correlations among molecular traits across all omics layers to explore DNA methylation - gene expression - protein - metabolite regulatory pathways. For example, we found that DNA methylation of gene *PCR1* at cg01407062 may influence lung cancer development by regulating the expression of *ANKRD35*, the protein fructose-bisphosphate aldolase B, and subsequently fatty acids. Protein fructose-bisphosphate aldolase B was positively correlated with lung cancer-associated fatty acids, including heptanoic acid, pelargonic acid, and caprylic acid. As a member of the Aldolase family, fructose-bisphosphate aldolase B catalyzes reactions in glycolysis and has been shown to regulate malignancy progression through various signaling pathways, such as PI3K/AKT/mTOR, GSK-3β, and Wnt^53–55^. Deregulation of the mTOR pathway has been linked to tumorigenesis and malignant progression in NSCLC^56^. Gene expression of *ANKRD35* was positively correlated with fructose-bisphosphate aldolase B. The Kyoto Encyclopedia of Genes and Genomes (KEGG) enrichment analysis indicated that knockdown of *ANKRD35* can activate many types of pathways, such as renal cell carcinoma, EGFR tyrosine kinase inhibitor resistance, and the mTOR signaling pathways^57^. Furthermore, *PRC1*, an interactor with *ANKRD35*, whose DNA methylation sites (cg01407062) were negatively associated with the expression *ANKRD35* (recorded in BioGRID). Finally, this analysis recovered results of investigating potential molecular mechanisms of lung cancer. The cg07493874-expression of *CLPTM1L* regulatory pathway was previously identified in NSCLC^16^. We expanded the exploration of potential molecular pathways to include additional omics layers. In total, we identified 106 potential regulatory pathways associated with lung cancer.

Our study has several strengths. First, to our knowledge, this is the first and the most comprehensive xWAS of lung cancer by systematically integrating alliance-based summary-level xQTL data from multiple omics layers with consortium-based individual-level GWAS data among the European-ancestry population. A systematic study of biomarkers in all omics layers by using the lung cancer consortium resources with the largest sample size provides us a unique opportunity to identify novel genes.

Second, to ensure the robustness of the results, we adopted a rigorous two-phase analytic strategy. To detect biomarkers with weak-to-moderate effect sizes in overall population and subgroup-specific biomarkers in subgroup populations, we meta-analyzed all independent participants from OncoArray, TRICL, PLCO and UKB. Additionally, biomarkers with strong effect sizes significant in all populations were also summarized. Finally, we released a free online platform, LungCancer-xWAS. We aim to provide comprehensive descriptions of signals in different scenarios, and offer more evidence for downstream biological studies in future. Third, it is increasingly evident that different molecular traits interact rather than act in isolation to influence lung cancer. To uncover potential mechanisms, we performed multi-omics integrative analyses to explore how these interactions impact lung cancer, and discovered over hundreds of across-layer pathways. Our results highlight that integrating xQTL summary-level data from multiple omics layers along with GWAS data can help deepen our understanding of disease molecular mechanisms.

There are still several limitations. First, developing and measuring the predictive performance of models typically require individual-level data and independent validation sets, but we lacked publicly available xQTL individual data. However, the LDpred2 ‘*auto*’ option we used enables prediction model development without requirement of validation sets, and we assessed model performance using *h*^2^ values for molecular traits. Second, the correlations among molecular traits in potential regulatory pathways provide data-driven evidence but may be influenced by confounding factors and reverse causality, warranting further mechanistic studies. Finally, incorporating other available xQTL data, such as splicing, histone, and chromatin accessibility QTL, could offer more possibility into novel lung cancer risk genes and pathogenic pathways.

## Methods

### Data Resource and Study Population

#### Lung Cancer GWAS Data

##### ILCCO-OncoArray

The ILCCO-OncoArray was established to enhance the understanding of the genetic architecture of common cancers^6^. The OncoArray GWAS was initially designed to analyze the genotype information of 57,775 individuals, sourced from 29 studies conducted across North America, Europe, and Asia. Informed consent was obtained from all participants. The studies received approval from local internal review boards or ethics committees and were conducted under the supervision of trained personnel. Genotyping of 517,820 SNPs in ILCCO-OncoArray was completed at the Center for Inherited Disease Research, the Beijing Genome Institute, the Helmholtz Zentrum München, Copenhagen University Hospital, and the University of Cambridge in Illumina Infinium OncoArray platform. Before standard quality control (QC) of data, the intentionally duplicated samples and samples from unrelated OncoArray studies and HapMap control individuals of European, African, Chinese, and Japanese origins were removed.

##### TRICL

The TRICL Research Team is part of the Genetic Associations and MEchanisms in ONcology (GAME-ON) Consortium^58^. All participants provided written informed consent. All studies were reviewed and approved by institutional ethics review committees at the involved institutions. The TRICL GWAS was originally designed to profile the genotype information of 12,651 participants. The genotype data of TRICL were generated from the Affymetrix Axiom Array, which contained 404,740 single nucleotide polymorphisms (SNPs).

##### PLCO

The PLCO Cancer Screening Trial is a large population-based randomized trial, initiated and funded by the National Cancer Institute (NCI). PLCO aimed at assessing the impact of screening on cancer-related mortality and secondary endpoints among more than 150,000 individuals aged 55 to 74^59^. Following the completion of the screening procedure in 2006, participants have been followed up for cancer incidence and mortality. The genotype data of PLCO were generated from the Illumina GSA (673,132 SNPs), OncoArray (474,276 SNPs), and historical data including Illumina OmniExpress (OmniX) (715,823 SNPs), Omni2.5M (Omni25) (2,310,570 SNPs) and Human Quad 610 (580,912 SNPs) arrays. The duplicated samples between different platforms were removed.

##### UKB

UK Biobank is a large-scale prospective study of individuals aged 40 to 70, who were assessed between 2006 and 2010 at various centers^60^. These participants underwent evaluations at assessment centers from 2006 to 2010. Blood samples were collected for genomic data, and participants completed questionnaires regarding their medical histories and environmental exposures. Various physical measurements were also taken. Lung cancer cases were defined by referencing the International Classification of Diseases, Tenth Revision (ICD-10) codes (field ID: 40006, 41202), as well as self-reported cancer diagnosis and histological types (field ID: 20001). Genetic data from the full UK Biobank, consisting of 488,377 individuals, were assayed using the Affymetrix UK BiLEVE and UK Biobank Axiom arrays.

Details of QC procedures and results were described in Supplementary Note and Supplementary Fig. 14.

#### xQTL Data Resource

##### Methylation QTL (meQTL)

The meQTL summary statistics were sourced from the Genetics of DNA Methylation Consortium (GoDMC)^17^, an international collaboration of human epidemiological studies aimed at investigating the genetic architecture of DNA methylation (DNAm) and exploring its potential role in disease mechanisms through large-scale epigenomic and genomic analyses. In total, 244,533 DNAm were measured in whole blood or cord blood using HumanMethylation450 or EPIC arrays in 27,750 European participants from 36 cohorts.

##### Expression QTL (eQTL)

The eQTL summary statistics were sourced from the eQTLGen Consortium^18^. Values from blood array expression data and RNA sequencing data were logarithmically transformed. After filtering, cis- and trans-expression quantitative trait locus (eQTL) analyses were performed using blood-derived 19,942 gene expressions from 31,684 European individuals from 38 different datasets.

##### Protein QTL (pQTL)

Plasma samples from 40,004 Icelanders were collected as part of two main projects^19^: the Icelandic Cancer Project, which accounted for 52% of the participants (with samples collected between 2001 and 2005), and various genetic programs at deCODE genetics, Reykjavík, Iceland (48%), which quantified 4,719 proteins in 35,559 Icelanders.

##### Metabolite QTL (metabQTL)

The metabQTL summary statistics were derived from the association results of a GWAS on the human metabolome conducted by So-Youn Shin et al^20^. Metabolites were profiled by liquid-phase chromatography coupled with tandem mass spectrometry (LC-MS) in peripheral plasma or serum to advance our understanding of the role of inherited variation in blood metabolic diversity. We obtained the association data from Metabolomics GWAS Server^20^, which contains all association statistics for 486 metabolites based on 7,824 European adults from the Twins UK and KORA (Cooperative Health Research in the Region of Augsburg) F4 studies.

The demographic characteristics and biomarker profiles of the four types of xQTL summary statistics were summarized in Supplementary Table 1 (a).

### Prediction Models Development of Molecular Traits

First, we developed molecular traits prediction models based on xQTL summary statistics. For each site, only SNPs with MAF > 0.01 were retained. We built statistical models using LDpred2 in the *bigsnpr* package of R to predict molecular traits.

LDpred2 is a widely used method for deriving polygenic risk score (PRS), which adopts a Bayesian approach for SNP selection based on summary statistics and takes into account linkage disequilibrium (LD) between the SNPs^61^. We used LDpred2 with the sparse automatic option, where the *sparse* option enables LDpred2 to learn effects equal to zero without sacrificing predictive accuracy, and where the *auto* option enables LDpred2 to automatically estimate the parameters of the model, namely sparsity *p* and the SNP heritability *h*^2^, without the need of validation data to tune hyper-parameters. We ran 30 Gibbs sampler chains for each molecular trait to verify the convergence of the model. We calculated the range of the *imputed* correlations between variants and molecular traits for each model, and models with correlation above their 95^th^ percentile were retained. The final model was randomly selected from the retained models for each site.

The *h*^2^_median_ values and *h*^2^_model_ values were calculated to assess the prediction performance of the models. The *h*^2^_median_ was defined as the median of the estimated heritability across all iterations of the retained models, while the *h*^2^_model_ represented the mean heritability of the selected models. Only models with satisfactory prediction performance, defined by (1) *h*^2^_median_ ≥ 0.01; (2) *h*^2^_model_ ≥ 0.01, were considered eligible.

### A Two-Phase Analysis of Associations between Genetically Predicted Molecular Traits and Lung Cancer

We adopted a two-phase (discovery and replication) analytic strategy to identify molecular traits associated with lung cancer. In the discovery phase, we used prediction models of molecular traits developed by LDpred2 and genotypes from lung cancer GWAS datasets to impute biomolecule levels for each individual. Logistic regression models were then applied to test the associations between genetically determined molecular traits and lung cancer within ILCCO-OncoArray, adjusted for age, sex, smoking status, platform (for PLCO only), and the top 10 principal components to account for potential population structure. The FDR was set to be 0.05 using Benjamini-Hochberg method^62^ to correct for multiple testing. In the replication phase, to validate the significant signals identified in the discovery phase, fixed-effect meta-analyses using independent datasets from TRICL, PLCO, and the UK Biobank were performed. Molecular traits were considered significant if they met the following criteria: (1) *q*-FDR < 0.05 in the discovery phase; (2) *P*_meta_ < 0.05 in the replication phase; (3) a consistent effect direction across both phases.

To nominate novel genes for lung cancer, we performed annotation for the CpGs and metabolites. For epigenomics, we used annotation obtained from the Illumina 450K platform (GEO: GPL18809) to annotate the closest genes and regions of the CpGs. For metabolomics, we used metabolite-associated loci from metabQTL to annotate the genes and the regions for metabolites^20^. We determine novel biomarkers and genes if they were never reported their lung cancer susceptibility before. We searched and synthesized genes have been nominated by previous studies for different omics layers. For genomics layer, we systematically searched publicly available studies in the GWAS Catalog (see URLs), setting the significance threshold for SNPs at *P* < 5 × 10^-8^. We excluded studies with sample sizes below 1,000 and those with non-European ancestry participants. For genetic variant selection, SNPs located in intronic regions were removed. In total, six studies with 57 SNPs across 77 genes were retained as previous GWAS-nominated genes. For epigenomics and transcriptomics layer, we cross-referenced reported lung cancer risk related CpGs and genes from the EWAS Atlas^21^ and TWAS Atlas^22^. For proteomics and metabolomic layer, we conducted searches on PubMed using the phrases *lung cancer* combined with *protein* or *metabolite*. We then removed studies that included samples not of European ancestry as well as those that used biomarkers not derived from blood.

### Mendelian Randomization

A two-sample MR was performed to further validate the robustness of the associations identified using the two-phase strategy in xWAS as part of the sensitivity analyses ^11^. For a more detailed description of the MR set-up, see the Supplementary Note. We are not to assert causal relationships between the molecular traits and lung cancer as some of the key assumptions of MR may not follow strictly. Nevertheless, the findings from MR can serve as a valuable complement to the associations identified from xWAS, offering evidence for the consistency of our strategies and the findings^11^.

### Meta-Analyses of Associations between Genetically Predicted Molecular Traits and Lung Cancer among Overall Population and Different Subgroups

To increase statistical power and improve the detection of novel signals in both general populations and specific subgroups, we conducted a series of meta-analyses. Estimates from the ILCCO-OncoArray, TRICL, PLCO, and UK Biobank cohorts were combined using a fixed-effect model for the overall population as well as for various subgroups, including NSCLC, SCC, LUAD, LUSC, males, females, ever smokers, and never smokers. Multiple testing correction was applied using the Benjamini-Hochberg method, setting the FDR at 0.05.

### Multi-omics Integrative Analyses

Lung cancer associated biomarkers identified by our xWAS were mapped to genes based on their physical positions. We aimed to depict lung cancer genes overlapped different omics layers. Genes were nominated as overlapping genes across different omics layers if meeting the following criteria: (1) they were nominated as significant genes by the two-phase analytic strategy in any one of layers or by previous GWASs, and (2) they showed nominal significance in the meta-analysis of all datasets (*P* < 0.05) in at least one of other omics layers.

In order to explore the cross-layer pathways, we adopt two methods. (1) For these nominated cross-layer overlapping genes, we evaluated pairwise correlation between two biomarkers from different omics layers. Significant correlations among these biomarkers mapping to a same gene indicated there was a across-layer pathway which dominantly driven by the gene from upstream to downstream omics layers. (2) Metabolites, as terminal ends, play a crucial role in lung carcinogenesis. Therefore, we utilized a hill-climbing strategy from downstream to upstream, to detect pathways consisting of multiple genes. First, we begun with lung cancer associated metabolites, and then determined their significantly correlated proteins in upstream. Next, genes whose expressions significantly correlated with proteins were established. Finally, DNA methylation CpG probes correlated with genes were pinpointed. Pearson correlation tests were used and multiple testing were corrected by FDR method.

#### Statistical analysis

Quality control of lung cancer GWAS data was conducted using PLINK 2.0 and imputed with TopMed. Polygenic risk scores were calculated using the LDpred2 method in the R package *bigsnpr*. Mendelian randomization (MR) estimates were generated with the *TwoSampleMR* package. Association analysis was performed with the *glm* function in R, and additional statistical analyses were completed in R v4.3.0.

## Data availability

Access URLs for four lung cancer GWAS individual data are as following:

- ILCCO-Oncoarray data are available from https://www.ncbi.nlm.nih.gov/projects/gap/cgi-bin/study.cgi?study_id=phs001273.v3.p2.
- TRICL data are available from https://www.ncbi.nlm.nih.gov/projects/gap/cgi-bin/study.cgi?study_id=phs001681.v1.p1.
- PLCO data are available from https://www.ncbi.nlm.nih.gov/projects/gap/cgi-bin/study.cgi-study_id=phs001286.v2.p2.
- UK Biobank data are available from https://www.ukbiobank.ac.uk/.

Access URLs for four types of xQTL summary statistics as following:

- methylation QTL statistics are available from http://mqtldb.godmc.org.uk/. Expression QTL statistics are available from: www.eqtlgen.org.
- Protein QTL statistics are available from: https://www.decode.com/.
- Metabolite QTL statistics are available from http://metabolomics.helmholtz-muenchen.de/gwas.

The following are the URLs of the public data resources used in this article:

- GWAS catalogue: https://www.ebi.ac.uk/gwas/search?query=lung%20cancer#association.
- GeneCards: https://www.genecards.org.
- BioGRID: https://thebiogrid.org/114517/summary/homo-sapiens/prc1.html.

The following are the URLs of the software used in this article:

- PLINK 2.0: https://www.cog-genomics.org/plink/.
- TopMed: https://imputation.biodatacatalyst.nhlbi.nih.gov/#!.
- R v4.3.0: https://www.r-project.org/.

## Acknowledgements

This study was supported by the National Natural Science Foundation of China (82220108002 to F.C., 82273737 to R.Z., 82473728 to Y.W.W., 82173620 and 82373690 to Y.Z.), the US National Institutes of Health (CA209414, CA249096, CA092824, and ES000002 to D.C.C., CA 249096 and CA209414 to Y.L.), Priority Academic Program Development of Jiangsu Higher Education Institutions (PAPD). R.Z. was partially supported by the Outstanding Young Teachers Training Program of Nanjing Medical University. We thank the patients and investigators who participated in OncoArray, TRICL, PLCO and UK Biobank for providing the GWAS data. We also thank all the QTL cohorts in the present work for making the statistics publicly available and are grateful to all the investigators and participants who contributed to those studies, including GoDMC for meQTL, FHS, LIFE Heart, LIFE Adult, NTR-NESDA, Young Finns Study, BEST, Fehrmann and Lifelines Deep for eQTL, the Icelandic Cancer Project and deCODE for pQTL, KORA F4 and TwinsUK for metabQTL.

## Author contributions

R.Z., Y. Wei, F.C., D.C.C., Y.L., C.I.A. ad R.J.H. were responsible for study conception and study design; Y.Z., Z.H. and H.S. critically reviewed the manuscript and results; Xiang Wang and Xuan Wang validated all analytical procedures and results; J.Z. developed the online platform; Y. Wang, A.W., N.X., X.X., M.Z. and Xuan Wang performed data analyses and interpretation; Y. Wang, A.W. and R.Z. wrote the draft. All authors approved the final version of the manuscript.

## Competing interests

The authors declare no conflict of interest.

## Notes

### Competing Interest Statement

The authors have declared no competing interest.

### Funding Statement

Funding: This study was funded by the National Natural Science Foundation of China (81530088 to F.C., 82273737 to R.Z., 82373690 to Y.Z., and 81973142 to Y.W.), US National Institutes of Health (CA209414, HL060710 and ES000002 to D.C.C.) and Priority Academic Program Development of Jiangsu Higher Education Institutions (PAPD). R.Z. was partially supported by the Qing Lan Project of the Higher Education Institutions of Jiangsu Province and the Outstanding Young Level Academic Leadership Training Program of Nanjing Medical University.
*Correspondence: Ruyang Zhang; Feng Chen; Yang Zhao; Yongyue Wei; David C Christiani.

### Author Declarations

Ethics committee of Nanjing Medical University gave ethical approval for this work.

