## Supplementary Note; Supplementary Fig. for "An integrative analysis of consortium-based multi-omics QTL and genome-wide association study data uncovers new biomarkers for lung cancer"

**Table of Contents**

Supplementary Note	1

Quality Control of GWAS Data	1

Two-sample Mendelian Randomization Model Set-up	2

Supplementary Figures	3

S1. Distribution of heritability values of molecular traits for four omics layers.	3

S2. Q-Q plots and heatmap for EWAS	4

S3. Q-Q plots and heatmap for TWAS.	5

S4. Q-Q plots and heatmap for PWAS.	6

S5. Q-Q plots and heatmap for MWAS.	7

S6. xWAS results in NSCLC population.	8

S7. xWAS results in SCC population.	9

S8. xWAS results in ever-smoking population.	10

S9. xWAS results in never-smoking population.	11

S10. xWAS results in male population.	12

S11. xWAS results in female population.	13

S12. xWAS results in LUAD population.	14

S13. xWAS results in LUSC population.	15

S14. Flowchart for QC of GWAS data	16

Supplementary References	17

### Supplementary Note

#### Quality Control of GWAS Data

***ILCCO-OncoArray,*** ***TRICL and UKB*** For these lung cancer GWAS datasets, we excluded those who were sex inconsistency, had low-quality DNA (call rate < 95%), or abnormal heterozygosity (deviation of ± 6SD), or were second-degree relatives or closer having identity by descent. SNPs were removed if meeting any of the following criteria: (1) sex chromosome, (2) call rate less than 95%, (3) Hardy-Weinberg equilibrium (HWE) test *P* less than 1.00 × 10^-12^, and (4) minor allele frequency (MAF) less than 0.001.

To estimate missing genotype information, we performed an imputation on the TOPMed online imputation server^[1]^, which phased haplotypes with Eagle v2.451 using TOPMed data (Version r2) as a reference panel that included 97,256 reference samples and 308,107,085 genetic variants^[2]^. The server performed imputations for ILCCO-OncoArray, TRICL, PLCO and UK Biobank using Minimac (version 4) software.

***PLCO*** Quality control (QC) and imputation for PLCO data were completed by the PLCO Atlas genotyping project. Details of QC procedures were described in the previous study^[3]^. Briefly, SNPs were removed if meeting any of the following criteria: (1) sex chromosome, (2) call rate less than 95%, (3) Hardy-Weinberg equilibrium (HWE) test P less than 1.00 × 10^-6^, and (4) minor allele frequency less than 0.01. Further excluded were those were sex inconsistency, had low-quality DNA (call rate < 95%), or abnormal heterozygosity (absolute values from PLINK method-of-moments F coefficients greater than 0.2), or were second-degree relatives or closer having identity by descent. After QC, genotype imputation was also performed on the Michigan Imputation Server *by using* the TopMed reference panel*.*

After genotype imputation, we filtered variants with poor imputation quality score *R*^2^ < 0.3, or with minor allele frequency ≤ 0.01, or in the duplicated position. All genetic variants were lifted to GRCh38/hg38 coordinates for maintaining consistency.

#### Two-sample Mendelian Randomization Model Set-up

For each site, the set of SNPs chosen by the prediction model in the xWAS was clumped down to independent SNPs (*r*^2^ < 0.1 within a 1000 kb window using the 1000 Genomes CEU reference panel for LD estimation) with a *P* value threshold of 0.01 using PLINK as instruments. The MR analysis was conducted using the *TwoSampleMR* (version 0.5.0) package^[4]^, using the Wald ratio (“mr_wald_ratio”), inverse-variance-weighted (“mr_ivw”), and Egger regression (“mr_egger_regression”) methods. Briefly, the Wald ratio approach was used when only a single SNP was used as an instrument and IVW approach was performed when more than one SNP was available. We also applied Cochran’s Q test to estimate the potential heterogeneity of MR estimates (*P* < 0.05) and considered signals whose MR-Egger intercept *P* < 0.05 as influenced by horizontal pleiotropy. Effects of molecular traits with heterogeneity were estimated by IVW random-effects model and those with horizontal pleiotropy were estimated by MR-Egger. False discovery rate (FDR) using Benjamini-Hochberg method was set to be 0.05 in order to correct multiple testing.

### Supplementary Figures

#### S1. Distribution of heritability values of molecular traits for four omics layers.

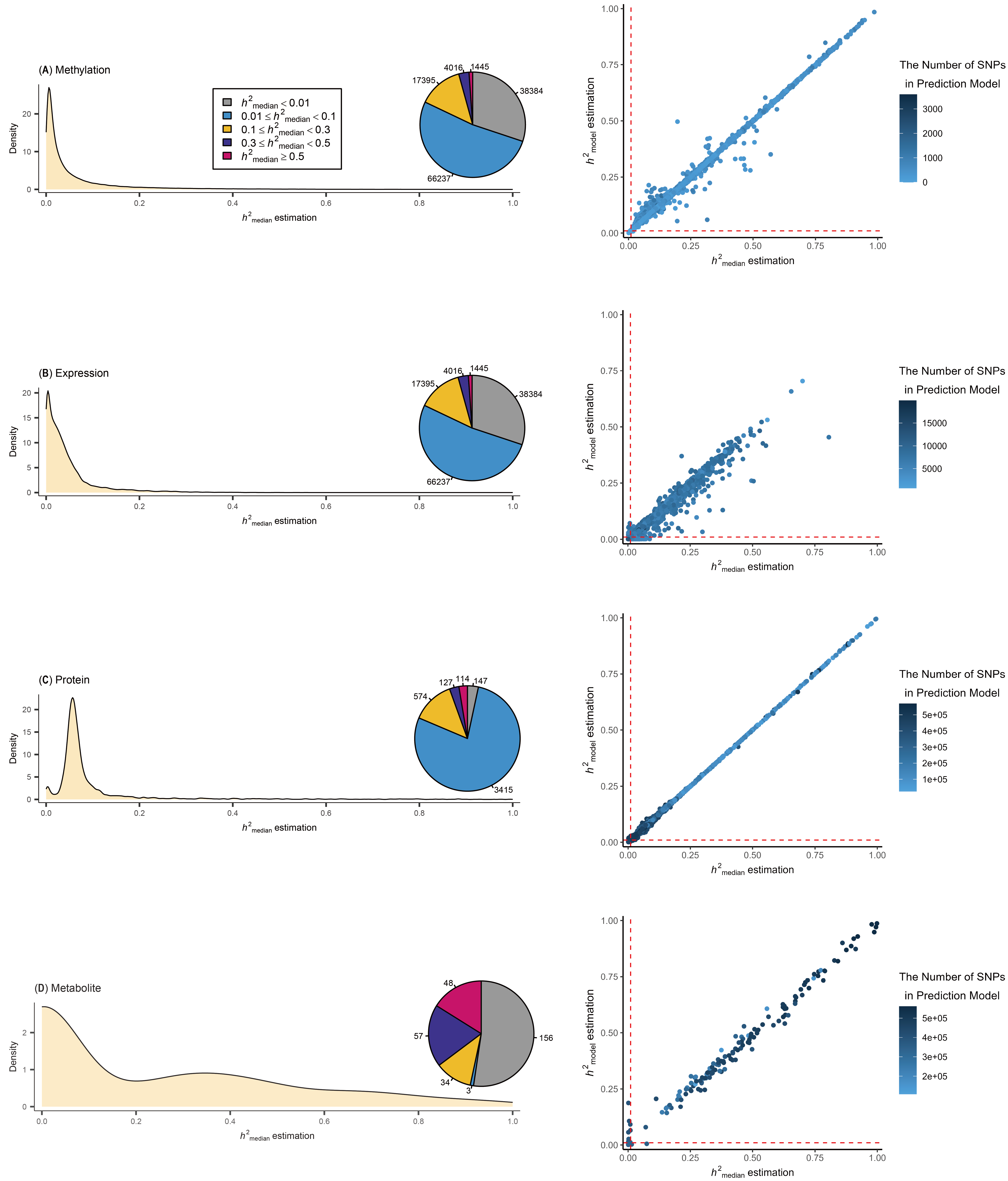

The *h^2^*_median_ estimated by LDpred2. Pie charts reflect the number of prediction models in a particular *h^2^*_median_ range (left). Prediction models with satisfactory prediction performance according to these criteria: (1) *h*^2^_median_ ≥ 0.01; (2) *h*^2^_model_ ≥ 0.01 (right).

#### S2. Q-Q plots and heatmap for EWAS

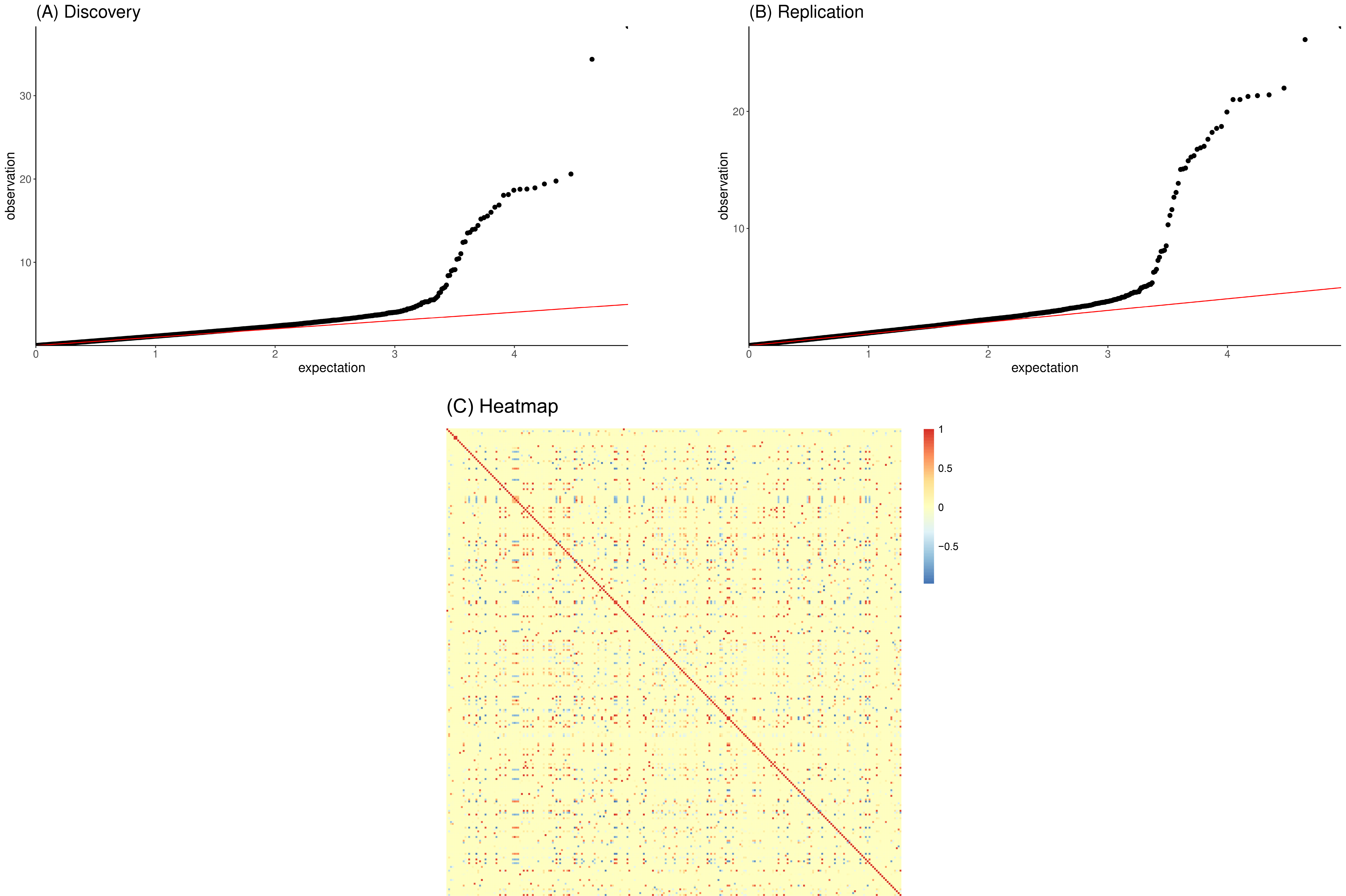

(A) Q-Q plot based on the discovery analysis, *λ* = 1.165; (B) Q-Q plot based on the replication analysis, *λ* = 1.071; (C) Heatmap showed correlation between CpG markers.

#### S3. Q-Q plots and heatmap for TWAS.

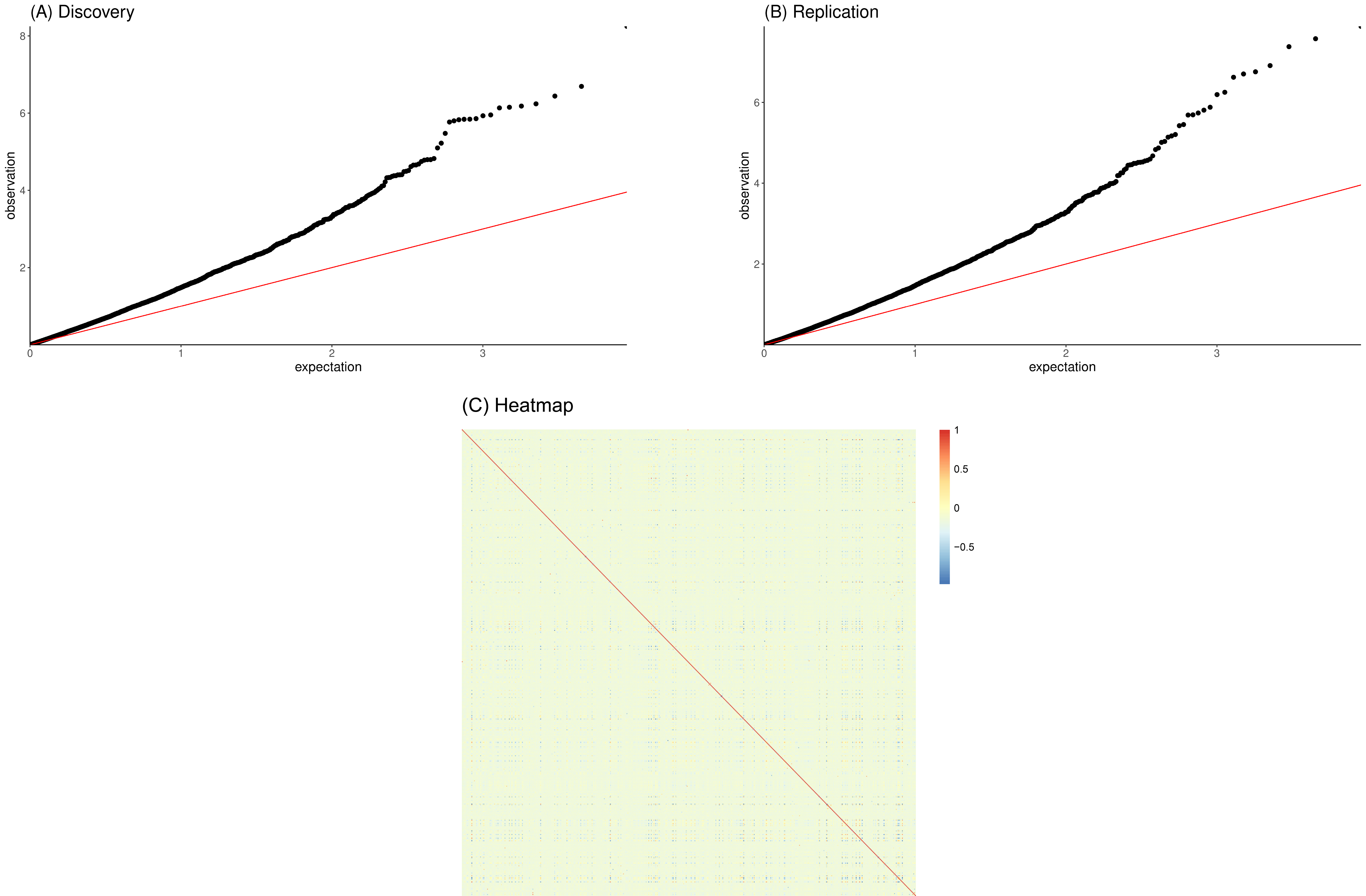

(A) Q-Q plot based on the discovery analysis, *λ* = 1.625; (B) Q-Q plot based on the replication analysis, *λ* = 1.527; (C) Heatmap showed correlation between gene expressions. Due to the strong correlation between many gene expressions, the genome inflation factor inflates to 1.63 and 1.53.

#### S4. Q-Q plots and heatmap for PWAS.

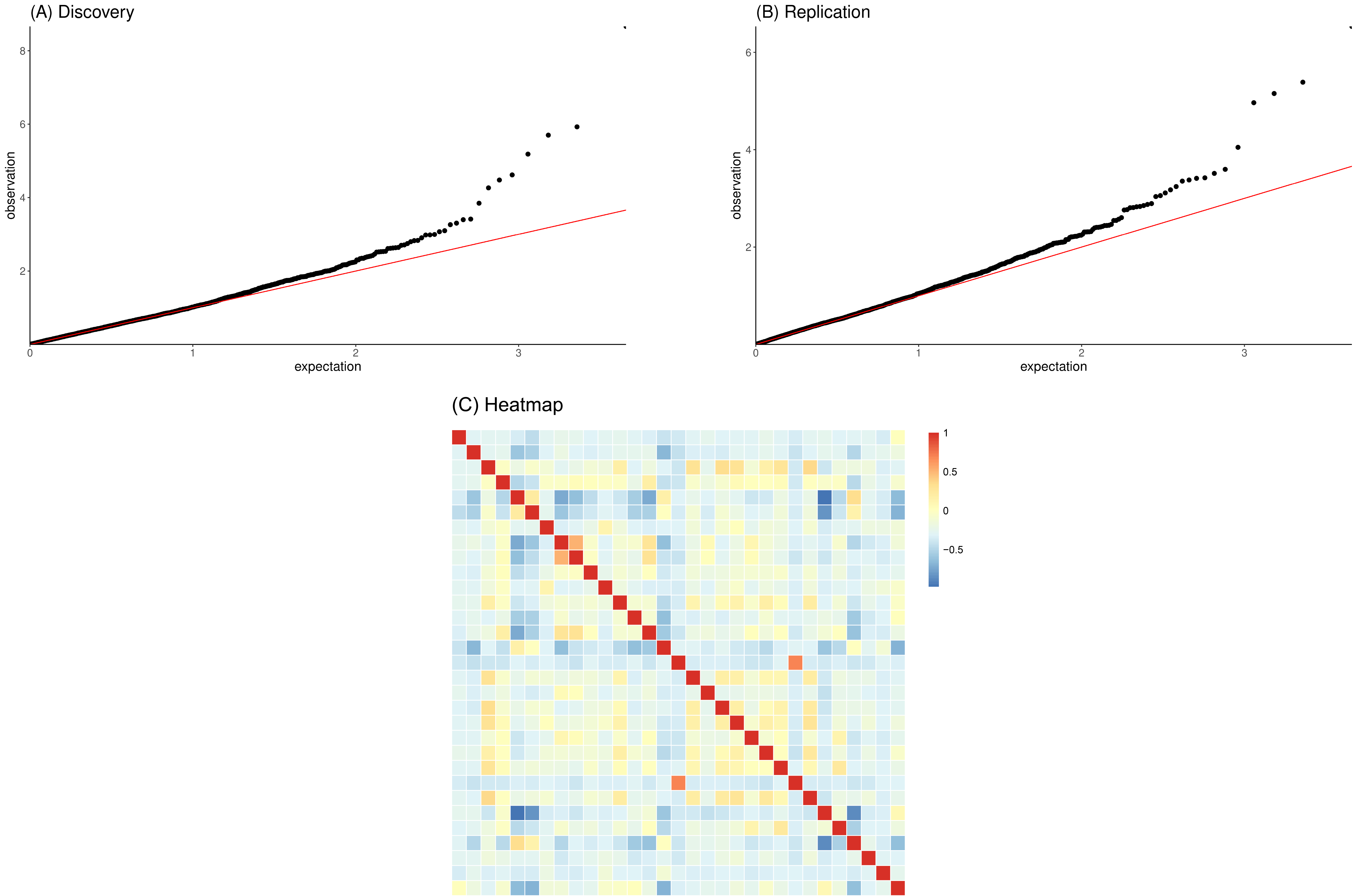

(A) Q-Q plot based on the discovery analysis, *λ* = 1.064; (B) Q-Q plot based on the replication analysis, *λ* = 1.101; (C) Heatmap showed correlation between proteins.

#### S5. Q-Q plots and heatmap for MWAS.

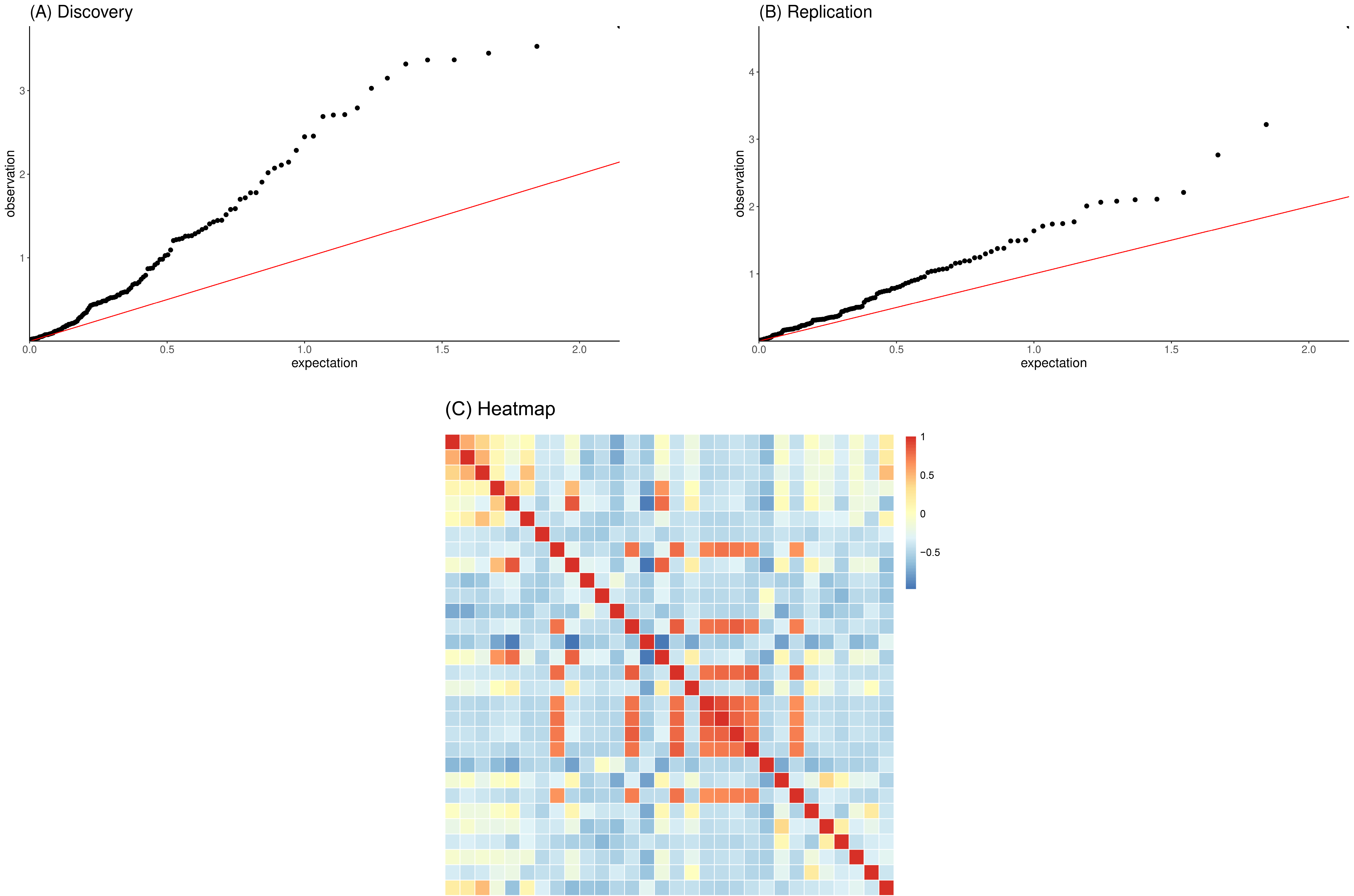

(A) Q-Q plot based on the discovery analysis, *λ* = 2.369; (B) Q-Q plot based on the replication analysis, *λ* = 1.662; (C) Heatmap showed correlation between metabolites. Due to the strong correlation between many metabolites, the genome inflation factor inflates to 2.37 and 1.67.

#### S6. xWAS results in NSCLC population.

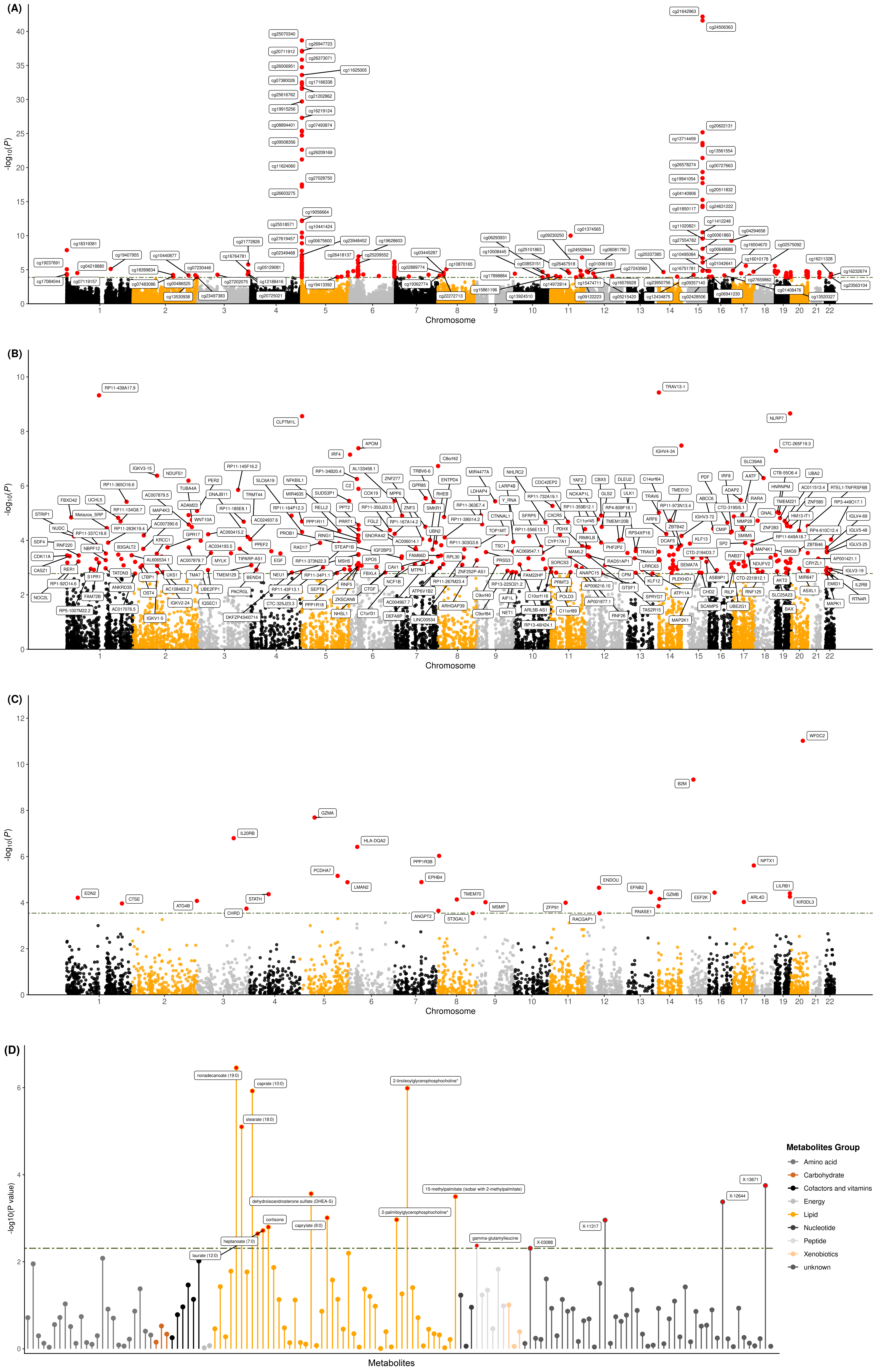

(A) Manhattan plot for DNA methylation analyses; (B) Manhattan plot for gene expression analyses; (C) Manhattan plot for protein analyses; (D) Lollipop plot for metabolite analyses. The green dotted line reflects the significant threshold of the *q*-FDR < 0.05 and is set at the highest unadjusted *P* value that is below that threshold. Each dot represents the genetically predicted molecular trait. The x axis represents the chromosome of the molecular traits in Manhattan plots of methylation, gene expression and protein, and represents metabolite in Lollipop plot of metabolite. The y axis represents the negative logarithm of the association *P* value.

#### S7. xWAS results in SCC population.

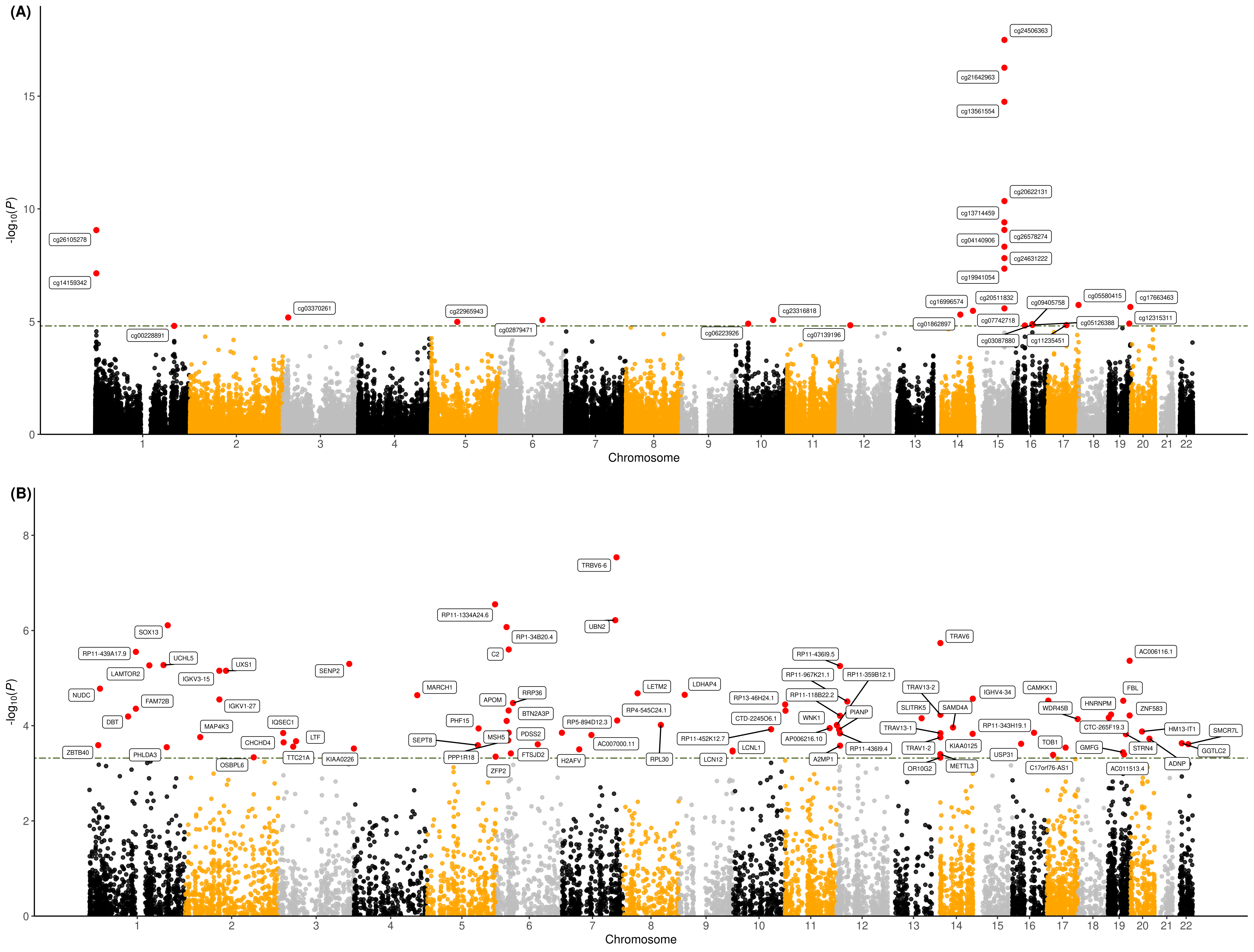

(A) Manhattan plot for DNA methylation analyses; (B) Manhattan plot for gene expression analyses. The green dotted line reflects the significant threshold of the *q*-FDR < 0.05 and is set at the highest unadjusted *P* value that is below that threshold. Each dot represents the genetically predicted molecular trait. The x axis represents the chromosome of the molecular traits in Manhattan plots of methylation and gene expression. The y axis represents the negative logarithm of the association *P* value. The red represents the molecular traits that pass the replication phase in the two-phase strategy.

#### S8. xWAS results in ever-smoking population.

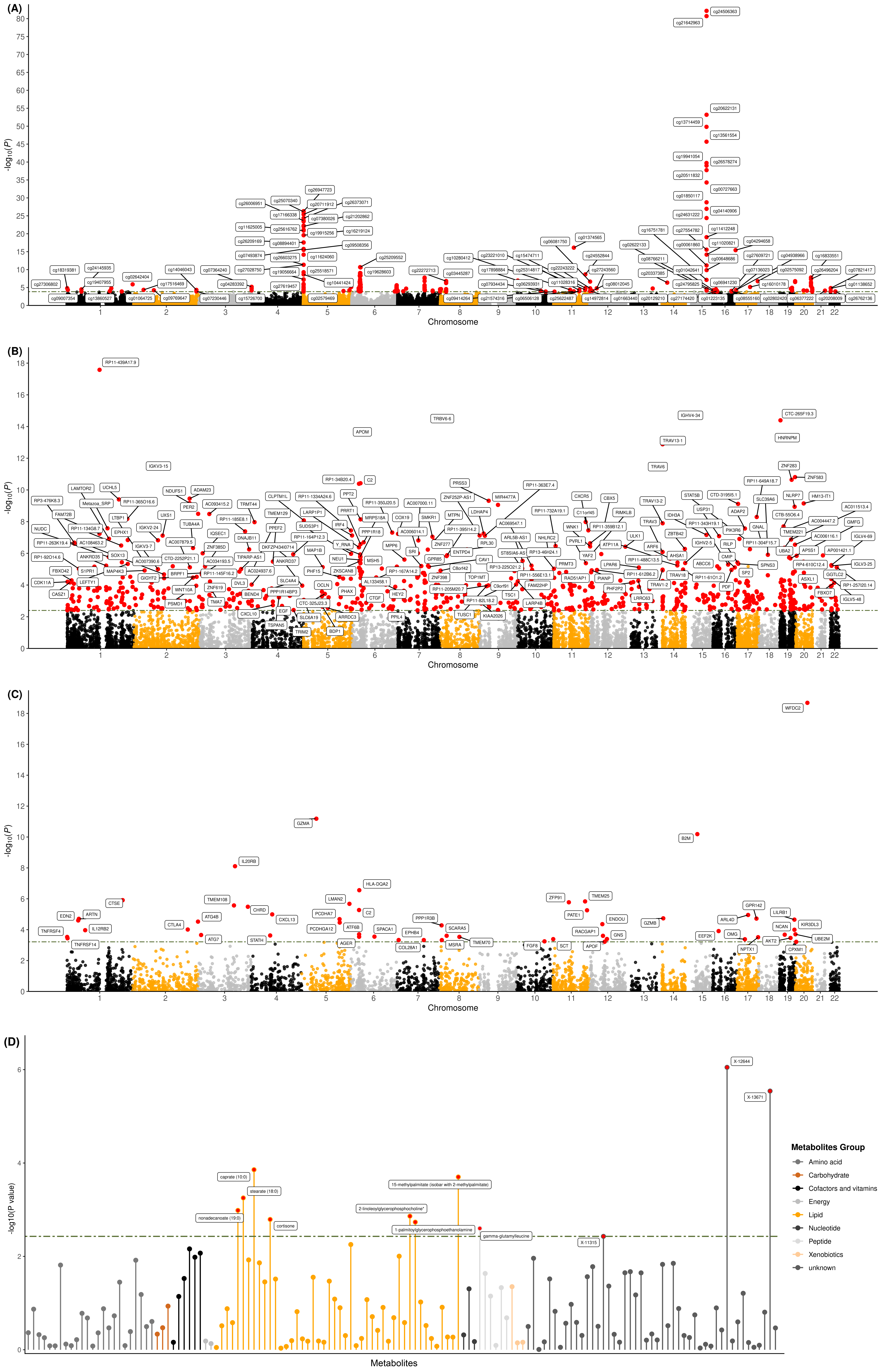

(A) Manhattan plot for DNA methylation analyses; (B) Manhattan plot for gene expression analyses; (C) Manhattan plot for protein analyses; (D) Lollipop plot for metabolite analyses. The green dotted line reflects the significant threshold of the *q*-FDR < 0.05 and is set at the highest unadjusted *P* value that is below that threshold. Each dot represents the genetically predicted molecular trait. The x axis represents the chromosome of the molecular traits in Manhattan plots of methylation, gene expression and protein, and represents metabolite in Lollipop plot of metabolite. The y axis represents the negative logarithm of the association *P* value.

#### S9. xWAS results in never-smoking population.

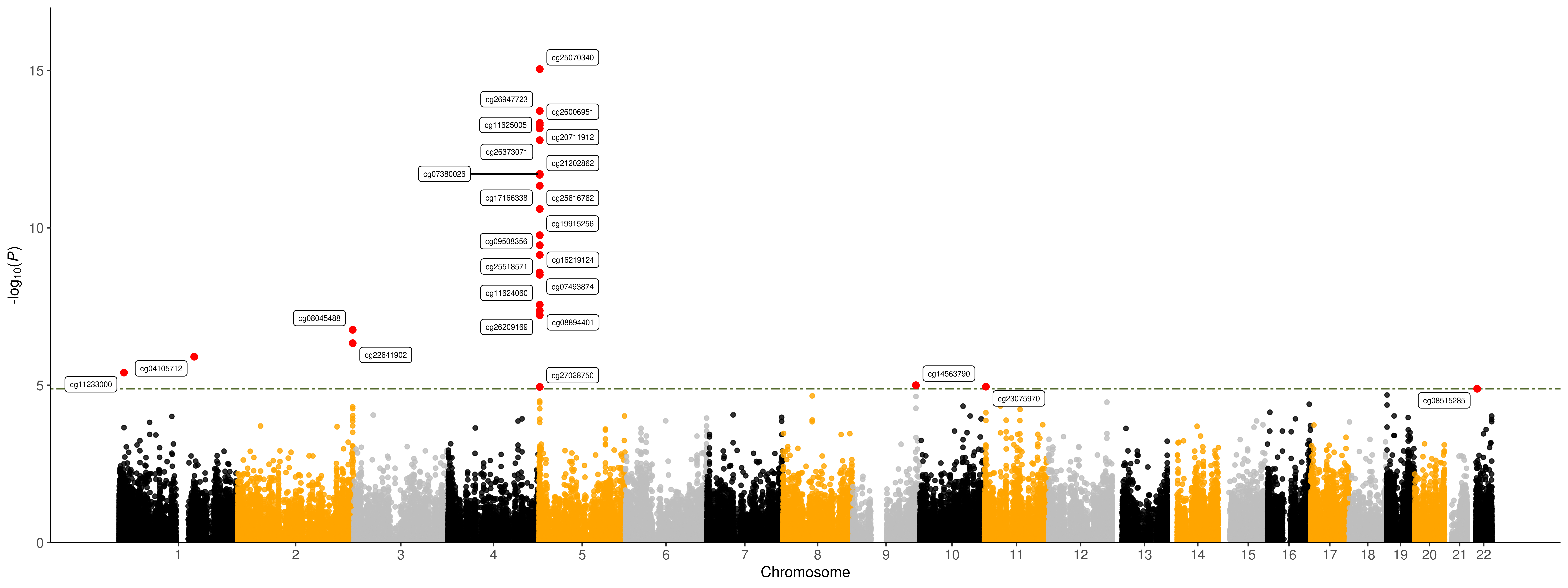

Manhattan plot for DNA methylation analyses. The green dotted line reflects the significant threshold of the *q*-FDR < 0.05 and is set at the highest unadjusted *P* value that is below that threshold. Each dot represents the genetically predicted DNA methylation marker. The x axis represents the chromosome of the DNA methylation markers in Manhattan plot. The y axis represents the negative logarithm of the association *P* value.

#### S10. xWAS results in male population.

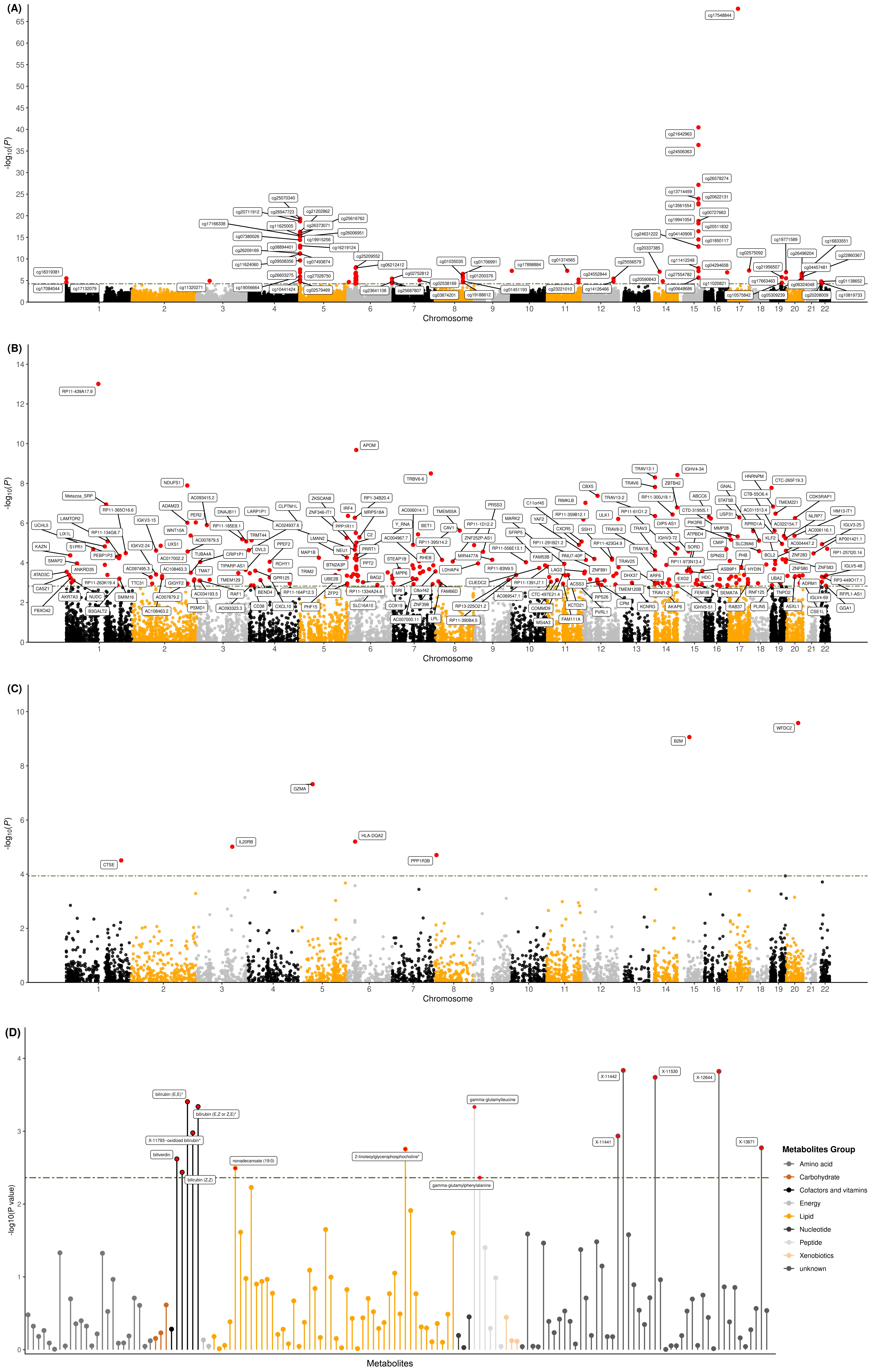

(A) Manhattan plot for DNA methylation analyses; (B) Manhattan plot for gene expression analyses; (C) Manhattan plot for protein analyses; (D) Lollipop plot for metabolite analyses. The green dotted line reflects the significant threshold of the *q*-FDR < 0.05 and is set at the highest unadjusted *P* value that is below that threshold. Each dot represents the genetically predicted molecular trait. The x axis represents the chromosome of the molecular traits in Manhattan plots of methylation, gene expression and protein, and represents metabolite in Lollipop plot of metabolite. The y axis represents the negative logarithm of the association *P* value.

#### S11. xWAS results in female population.

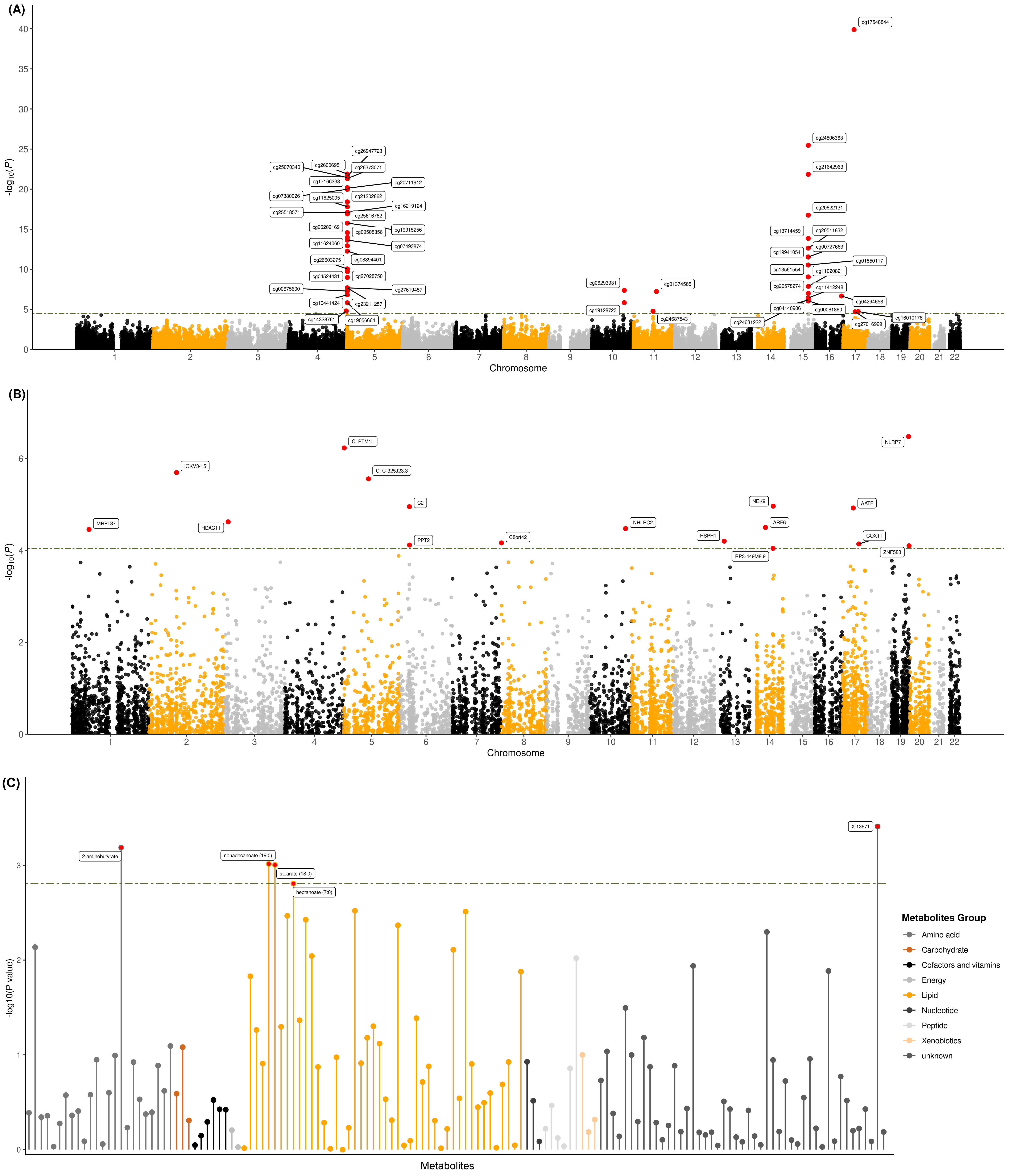

(A) Manhattan plot for DNA methylation analyses; (B) Manhattan plot for gene expression analyses; (C) Lollipop plot for metabolite analyses. The green dotted line reflects the significant threshold of the *q*-FDR < 0.05 and is set at the highest unadjusted *P* value that is below that threshold. Each dot represents the genetically predicted molecular trait. The x axis represents the chromosome of the molecular traits in Manhattan plots of methylation and gene expression, and represents metabolite in Lollipop plot of metabolite. The y axis represents the negative logarithm of the association *P* value.

#### S12. xWAS results in LUAD population.

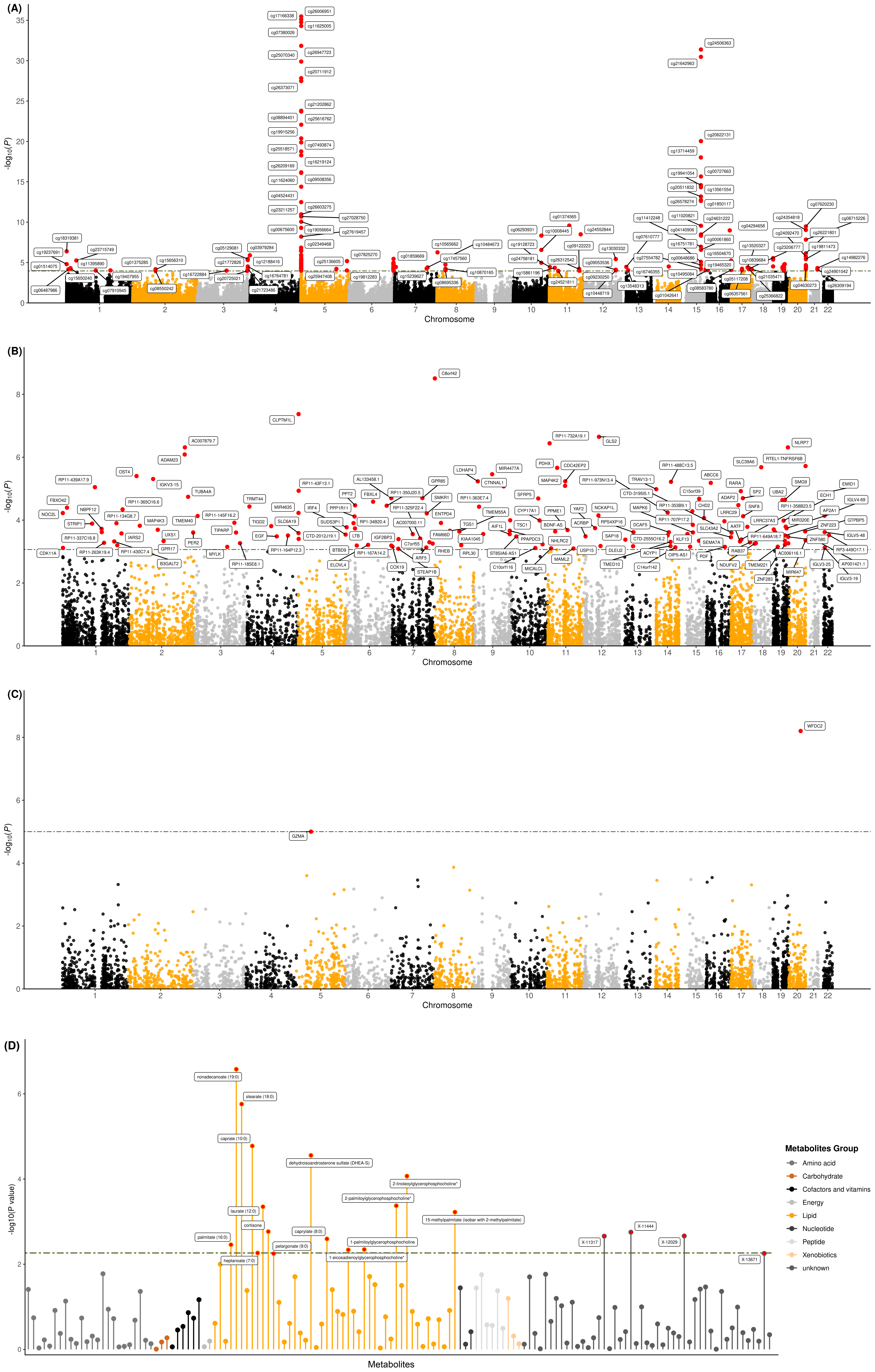

(A) Manhattan plot for DNA methylation analyses; (B) Manhattan plot for gene expression analyses; (C) Manhattan plot for protein analyses; (D) Lollipop plot for metabolite analyses. The green dotted line reflects the significant threshold of the *q*-FDR < 0.05 and is set at the highest unadjusted *P* value that is below that threshold. Each dot represents the genetically predicted molecular trait. The x axis represents the chromosome of the molecular traits in Manhattan plots of methylation, gene expression and protein, and represents metabolite in Lollipop plot of metabolite. The y axis represents the negative logarithm of the association *P* value.

#### S13. xWAS results in LUSC population.

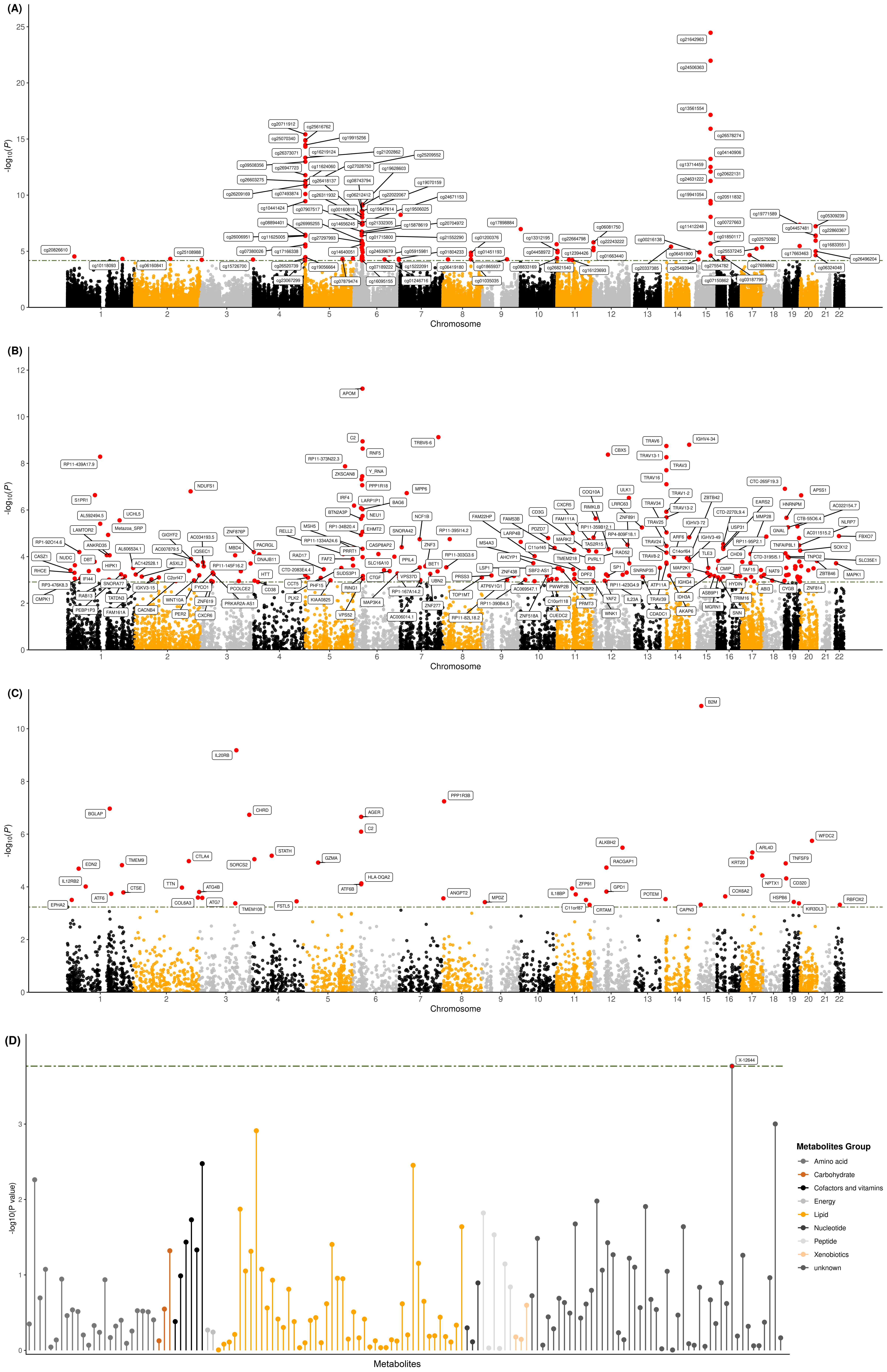

(A) Manhattan plot for DNA methylation analyses; (B) Manhattan plot for gene expression analyses; (C) Manhattan plot for protein analyses; (D) Lollipop plot for metabolite analyses. The green dotted line reflects the significant threshold of the *q*-FDR < 0.05 and is set at the highest unadjusted *P* value that is below that threshold. Each dot represents the genetically predicted molecular trait. The x axis represents the chromosome of the molecular traits in Manhattan plots of methylation, gene expression and protein, and represents metabolite in Lollipop plot of metabolite. The y axis represents the negative logarithm of the association *P* value.

#### S14. Flowchart for QC of GWAS data

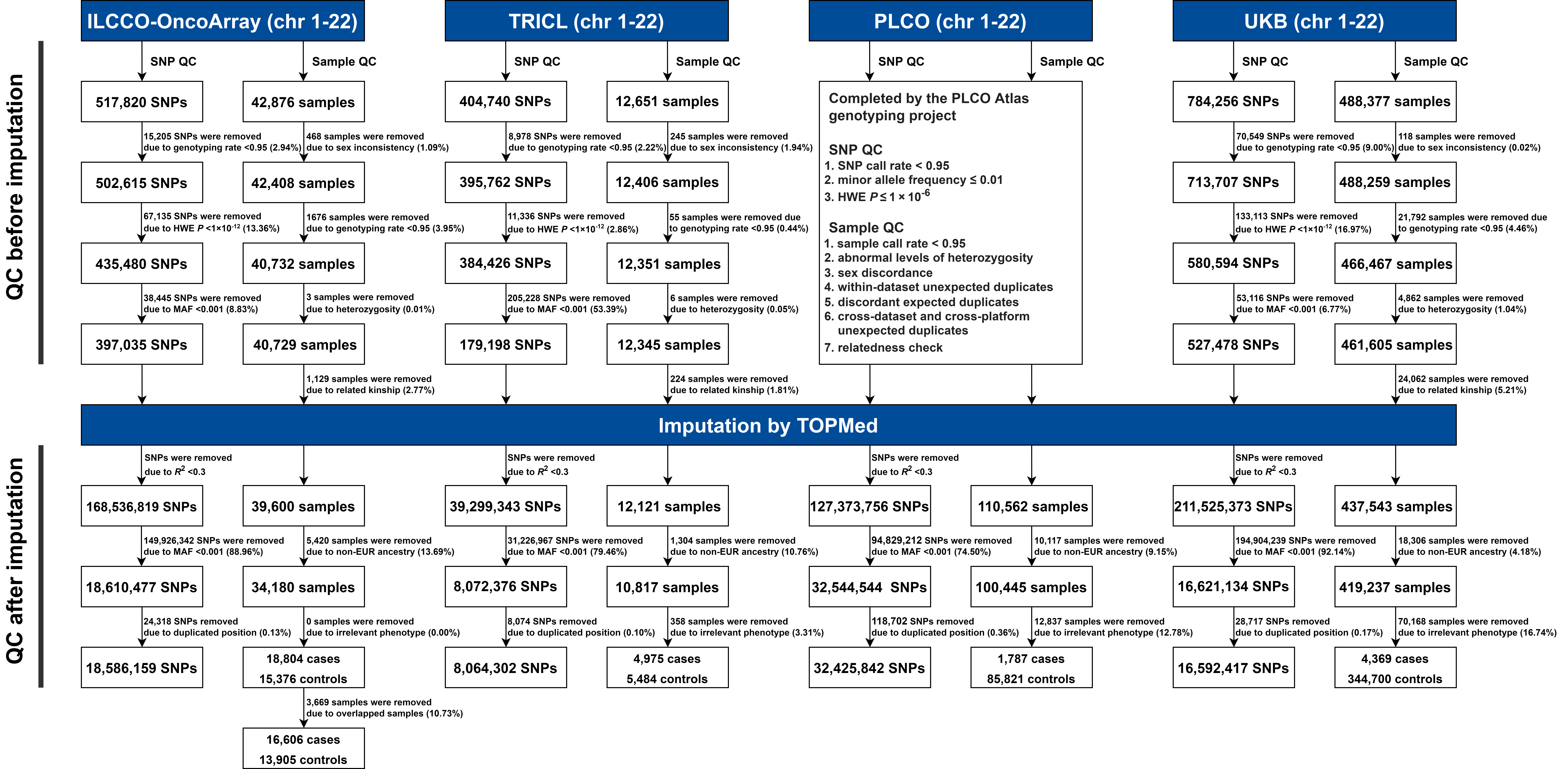
